# Learning lifetime disease liability reveals and removes genetic confounding in electronic health records

**DOI:** 10.64898/2026.02.15.26346336

**Authors:** Yazheng Di, Na Cai

## Abstract

Electronic health records (EHRs) have become the cornerstone of population-scale genetic studies^1^, but factors including patterns of healthcare use shape which and how diagnoses are recorded, leading to confounding effects in genetic associations with EHR codes^2^. In this study we propose EDGAR, a deep learning framework that recovers lifetime disease liability from EHR by aligning diagnostic codes with clinically validated measures and disease labels in a set of individuals prioritized through active learning. EDGAR yields representations that better capture disease-specific effects in genome-wide association analyses (GWAS). It also enables us to isolate a genetic factor that captures systemic biases in EHR codes, which distorts cross-disease correlations and drives spurious links with behavioral and socio-economic traits. We find that this factor generalizes across EHRs, and its identification in one EHR enables its removal from existing GWAS in another. Overall, our work presents a promising direction for improving specificity of EHR-based GWAS.

## Introduction

Genome-wide association studies (GWAS) are increasingly performed with data from electronic health records (EHR), transforming scale from a limiting factor^3–7^ into a defining feature of human genetics^1^. By linking genomic data to diagnostic codes, EHR-based cohorts^1^ or meta-analyses that rely heavily on them^8–10^ have now achieved sample sizes and GWAS statistical power that are nearly impossible to reach using detailed clinical assessments (deep phenotyping)^11,12^. This approach has essentially replaced proactively sampled deep phenotypes with passively collected diagnostic codes. However, many previous works^13–15^ including our own^16^ have shown that these “shallower” phenotypes yield non-specific genetic findings, and these effects are likely to propagate through analyses of disease etiology and cross-disease relationships^17–19^. There is therefore an urgent need for us to understand how systematic biases in EHRs influence genetic signals, and develop principled approaches for disentangling them from genetic effects on the underlying biological risk^2^.

How do EHR diagnostic codes reflect more than disease biology? We can think of an EHR diagnosis on an individual as reflecting a combination of their disease liability, their propensity to engage with the healthcare system^20–23^, and other operational reasons^24^. Healthcare engagement^25^ depends on behavioral traits such as care-seeking^26^ and interactions with clinical systems^27,28^, as well as structural disparities in healthcare access across sex, ancestry, and socioeconomic status^29–35^. Operational reasons include implicit bias in diagnoses^30,35,36^, heterogeneous institutional ICD coding practices^37^ and differences in incentives including billing-driven workflows^38,39^. These can distort records and introduce great heterogeneity within a single health system, as well as across them. Many of these factors are also heritable, collectively creating a landscape of systemic bias across diseases^40–47^. Additionally, if the underlying behavioral and operational drivers are preserved across healthcare systems^2,35^, the spurious genetic signal they produce would likely replicate across EHRs-based GWAS too, creating a ’circularity of bias’^15,17,48,49^ where systemic confounding can be mistaken for robust biological discovery.

The ideal phenotypes to use in genetic studies are lifetime disease liabilities that are free of systemic bias. This has motivated deep learning-based approaches aimed at inferring latent disease liability from EHR events, to break the ‘circularity of bias’. However, most existing models rely solely on EHR-derived labels for supervision and evaluation, and will therefore inevitably replicate systemic biases^50–54^. The hurdles are two-fold. First, independent anchors such as clinically-validated, detailed assessments of disease status (the deep phenotype^16^) are needed as supervision labels for model training. While these may also contain bias, they are unlikely to be affected by exactly the same systemic errors as EHR codes. This enables us to decouple true disease liability from method-specific noise^13,55^. However, these deep phenotypes are rarely recorded in EHRs^54^, and the budget to obtain them through patient call-back efforts (which are expensive and time-consuming) for targeted phenotyping^56–58^ is usually limited. Furthermore, how best to optimize label efficiency for learning lifetime disease liability, through prioritizing individuals to call back under a limited budget, has not been extensively explored.

Second, while predictions for disease liability may benefit from additional inputs in the form of biologically-valid, disease-relevant measures, and they are likely available in EHRs, there are technical challenges for integrating them with EHR codes for prediction tasks. On the one hand, current transformer-based prediction models^50–54^ are built upon a discrete-token paradigm^59^ optimized for clinical vocabularies (such as ICD codes), making them incompatible with high-dimensional continuous inputs like disease-relevant lab tests. For example, the recently published Delphi-2M^53^ model, which predicts future disease risks based on past EHR events, incorporates only a couple of quantitative measures as discretized tokens. On the other hand, tabular-based approaches^60,61^, though natively able to handle quantitative inputs, have not been shown to work on sparse EHR events.

To address these challenges, we propose a disease-liability prediction framework EDGAR (**E**HR **D**isease liability prediction for **G**enetic **A**rchitecture **R**ecovery), that aligns EHR records with deep phenotypes and disease-relevant measures. Recognizing that deep phenotypes are seldom recorded within routine EHRs and are costly to obtain, we augment our prediction framework with an active learning^62^ approach that prioritizes individuals for obtaining deep phenotypes, and demonstrate that this approach massively improves label efficiency. We apply these strategies to nine common diseases in the UKBiobank and perform GWAS and downstream analyses to evaluate the genetic relevance of the predicted liabilities. We then further utilize these predicted liabilities to isolate a genetic factor that accounts for systemic bias in the UKBiobank EHR, which distorts cross-disease correlations and drives spurious links with behavioral and socio-economic traits. Finally, we show that this EHR-systemic bias is generalizable and can be leveraged for bias removal from existing GWAS summary statistics in an external EHR, without requiring any model training on individual-level data.

Overall our findings demonstrate the feasibility, efficiency, and importance of lifetime disease liability prediction in EHR-based genetic studies, as well as the effectiveness of removing EHR-systemic bias at reducing spurious signals driven by socioeconomic and behavioural traits.

## Results

### Modelling of EHR codes in EDGAR

We focus on nine common diseases in 337,129 White British European individuals in UKBiobank (**Methods, Supplementary Methods**), including generalized anxiety disorder (GAD), major depressive disorder (MDD), anaemia (ANEM), chronic obstructive pulmonary disease (COPD), hyper-LDL-cholesterolemia (HLDL), non-alcoholic fatty liver disease (NAFLD), diabetes (DBT, inclusive of type 1 and type 2, see **Supplementary Methods**), hypertension (HTN), and osteoporosis (OST). For each of the nine diseases, we define three phenotypes (**Figure 1a**): (i) the binarized EHR phenotype is defined as the presence of a disease code from passively collected diagnosis codes in general practitioners (GP) and hospital inpatient records (HES, **Methods, Supplementary Table S1**); (ii) the deep phenotype is defined from diagnostic biomarkers or CIDI-adapted questionnaires, available in 8.50 - 97.27% of 337,129 assessed individuals in the dataset (**Methods**, **Supplementary Table S2**); and (iii) the predicted liabilities are from our deep learning framework. Hereafter, “phenotype” refers to these different phenotype definitions, whereas “disease” or “disorder” refers to the underlying condition.

**Figure 1.**
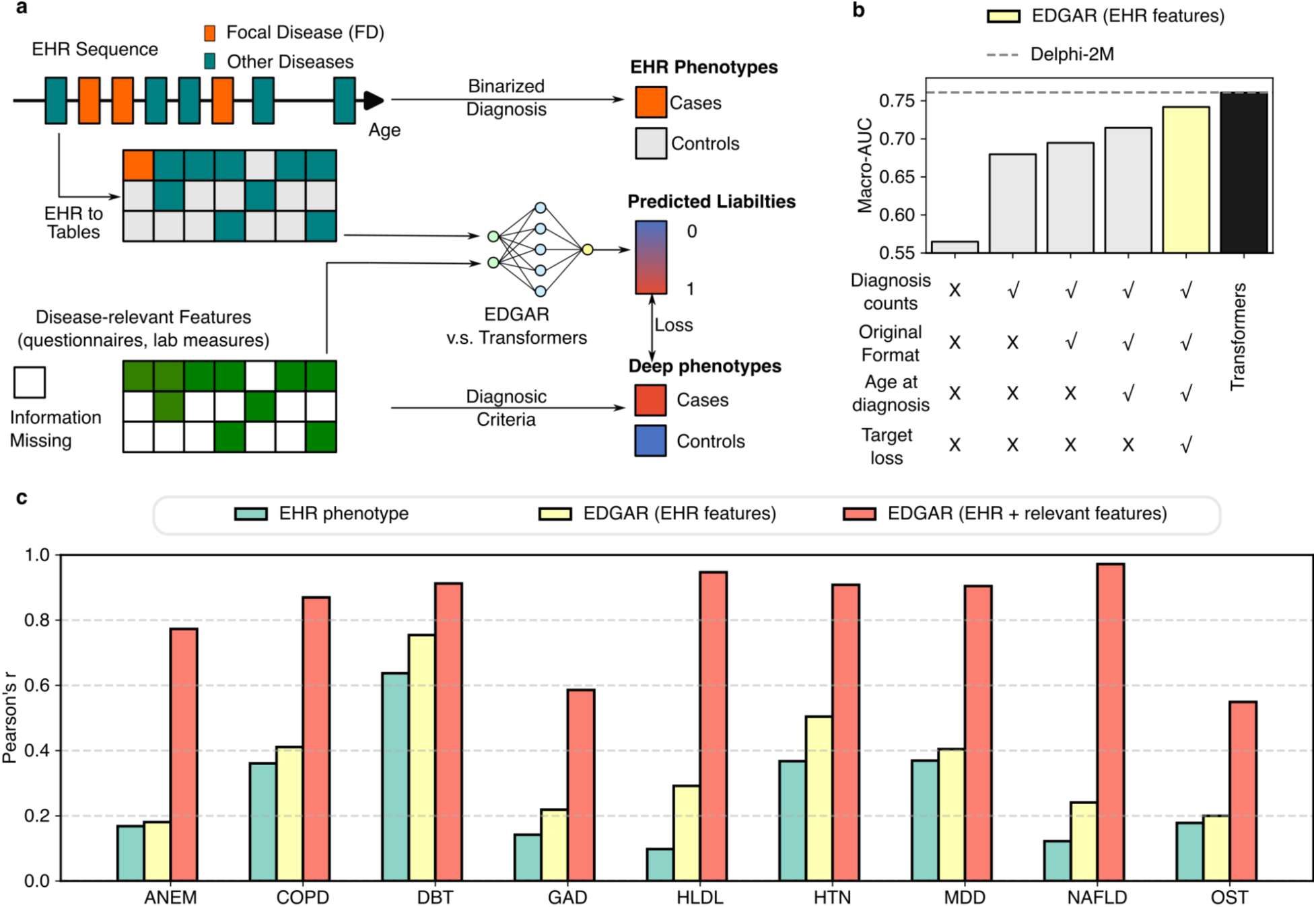
Phenotype definitions and disease liability prediction accuracy. **a)** Phenotype definitions for EHR phenotypes, deep phenotypes and predicted liability phenotypes in the EDGAR framework. **b)** Macro-AUC for models trained on EHR features only. Bars from left to right represent models of increasing complexity, differing by whether they use binary or counts of diagnosis codes; retain GP codes in their original Read2/CTV3 formats or map them to ICD-10; include age at diagnosis; and apply target loss. Gray bars denote baseline tabular models. The yellow bar shows our main tabular model EDGAR (same as in panel c), and the black bar indicates the transformer model trained on EHR sequences. The dashed line represents the published results from Delphi-2M (**Supplementary Table S3**), a transformer trained solely on EHR codes. It is included as an external anchor to provide context for state-of-the-art EHR modeling performance (**Supplementary Methods**), while our in-house transformer (black bar) ensures a direct architectural comparison under identical deep phenotype supervision. Disease-specific AUC is in **Extended Data** Figure 1a. **c)** Disease-specific accuracy (Pearson’s r) for EHR phenotype, EDGAR trained on EHR features only, and EDGAR trained on EHR with additional disease-relevant features.

We first assess predictiveness of EHR phenotypes for deep phenotypes of the corresponding diseases. We find that predictiveness ranges from low to modest (macro-AUC=0.64, Pearson’s r = 0.10–0.64, **Figure 1c**), and the only exception is DBT, where EHR phenotype achieves high consistency with the deep phenotype (AUC=0.94, r=0.64). The generally low predictiveness suggests EHR phenotype being a poor proxy for the true lifetime disease liabilities.

We then use our EDGAR framework to predict lifetime disease liability of nine common diseases (**Methods**). The EDGAR framework uses deep phenotypes as supervision labels, and counts of EHR codes available from both GP and HES records as tabular feature inputs that feed into an multi-layer perceptron (MLP) architecture adapted from AutoComplete (**Methods**). To identify the best input features obtainable from EHR events, we conducted a stepwise model augmentation analysis (**Figure 1b, Extended Data Figure 1a**). We find that (i) using the presence of diagnosis codes as binary features in the model does not yield predicted disease liabilities that are of improved predictiveness (macro-AUC = 0.56), consistent with EHR phenotype being poor proxy for disease liabilities; (ii) replacing binary diagnosis codes with the total counts per code substantially improves predictiveness (macro-AUC = 0.68), suggesting that the more times an individual has a code in the EHR for a disease, the more likely they are to have higher lifetime disease liability; (iii) retaining GP codes in their original format rather than mapping them to ICD-10 yields further gains (macro-AUC = 0.69), suggesting distinguishing between biases present in different data sources helps; (iv) incorporating diagnosis age information leads to additional improvements (macro-AUC = 0.71); and (v) using target-specific loss optimization (**Methods**) achieves the best overall performance (macro-AUC = 0.74).

We then ask whether there is an architectural bottleneck in our tabular model in EDGAR, by comparing it with transformer-based models^59,63^. We find that a transformer model trained on raw, sequential EHR codes, similar to the one deployed in a recently published model Delphi^53^ (**Supplementary Methods**), produces comparable predictive power (macro-AUC = 0.76, **Figure 1b, Extended Data Figure 1a**) to both the best tabular model as shown above (macro-AUC = 0.74) and Delphi itself (macro-AUC = 0.76, **Extended Data Figure 1a**, **Supplementary Table S3**). As Delphi is trained solely on EHR events for next event prediction, not on deep phenotype labels for lifetime liability prediction, it is not directly comparable with our EDGAR models (**Supplementary Methods**). Nevertheless, this suggests that our model is unlikely to gain further prediction accuracy by deploying more sophisticated deep learning models on solely EHR events.

## Improving accuracy using disease-relevant measures in EDGAR

We next proceed to add disease-relevant measures (available in 0.13 - 100% of 337,129 assessed individuals, **Supplementary Table S1 and Supplementary Table S4**) as features in our EDGAR model. We find that for almost all diseases tested, including disease-relevant features dramatically improves the performance of lifetime disease liability prediction (macro-AUC=0.98, Pearson’s r = 0.55-0.97, **Figure 1c, Extended Data Figure 1a**), where prediction accuracies for individual diseases are consistent with the availability of disease-relevant measures and deep phenotype labels (**Supplementary Methods**). We therefore use this full EDGAR model for the rest of the analyses presented in this paper.

To identify the features that most contribute to disease liability prediction for each of the nine diseases, we obtain SHAP^64,65^ values for our full EDGAR model (**Supplementary Methods**). For most diseases, the features with the highest SHAP values are the clinically validated diagnostic markers that, in combination with other criteria, define the disease (**Supplementary Figures S1-9**). To investigate if this causes data leakage and inflates prediction AUC, we perform targeted feature ablations to evaluate how the model behaves when primary diagnostic markers are unavailable (**Supplementary Methods)**. We find that this results in a limited decrease in prediction accuracy (e.g. ANEM: AUC 0.98→0.76; COPD: AUC 0.99→0.95; NAFLD: AUC 1.00→0.97), demonstrating the model’s ability to leverage the correlation between the input features that may capture biological redundancy, such that upon removal of a primary diagnostic marker, the model pivots to highly correlated features (e.g., AST in the absence of ALT for NAFLD, **Supplementary Tables S2,S5**).

## Evaluating feasibility of EDGAR under realistic data availability

While disease-relevant features dramatically improves prediction, it is likely they are available in far fewer individuals in routine EHRs that are not linked to biobanks like in UKBiobank. Similarly, all EDGAR predictions we show are performed with all available deep phenotype disease labels available in UKBiobank (up to 100% of assessed individuals). It is unlikely they are available at these rates in routine EHRs. As such, we simulate various scenarios when there are fewer individuals with deep phenotyped labels or disease-relevant measures.

We first simulate a realistic scenario in which disease-relevant measures are only available for patients who have interacted with the EHR system. To do this we restrict additional disease-relevant feature availability to individuals who have received a diagnosis for any of the nine diseases, resulting in a mean sample retention rate of 52.8% (SD = 4.5%, **Supplementary Table S4**). This simulation yields a decrease in prediction accuracy (macro-AUC = 0.88, Pearson’s r = 0.35–0.88; **Extended Data Figure 1b**), yet still outperforms models using EHR features alone. Further, using all available features (even those that are relevant for a different disease), in some cases, further improves model performance (macro-AUC: 0.97, Pearson’s r = 0.48-0.99, **Extended Data Figure 1b**), demonstrating features not conventionally deemed relevant to a disease may provide useful information for liability prediction. In other words, disease-relevant features in every available individual counts.

We then simulate scenarios where there are fewer individuals with deep phenotype labels. Here we assume deep phenotype labels can only be obtained through detailed assessments performed in patient call-back efforts: for every call-back budget for N individuals, we randomly select N individuals from the deep phenotype sample pool in UKBiobank to reveal their labels to train EDGAR. We find that increasing the labelled sample size N gives diminishing returns in prediction accuracy (variance explained R^2^ in the test dataset, **Figure 2b**), with N=20,000 (macro-AUC=0.97) reaching a similar performance as using all available deep phenotype data (macro-AUC=0.98, labelled N = 28,665 - 327,916, **Figure 1c**). This indicates that we need much fewer labelled individuals than available in UKBiobank, which suggests obtaining few deeply phenotyped individuals call-back studies may enable high quality EDGAR predictions of disease liabilities that are suitable for genetic studies.

**Figure 2.**
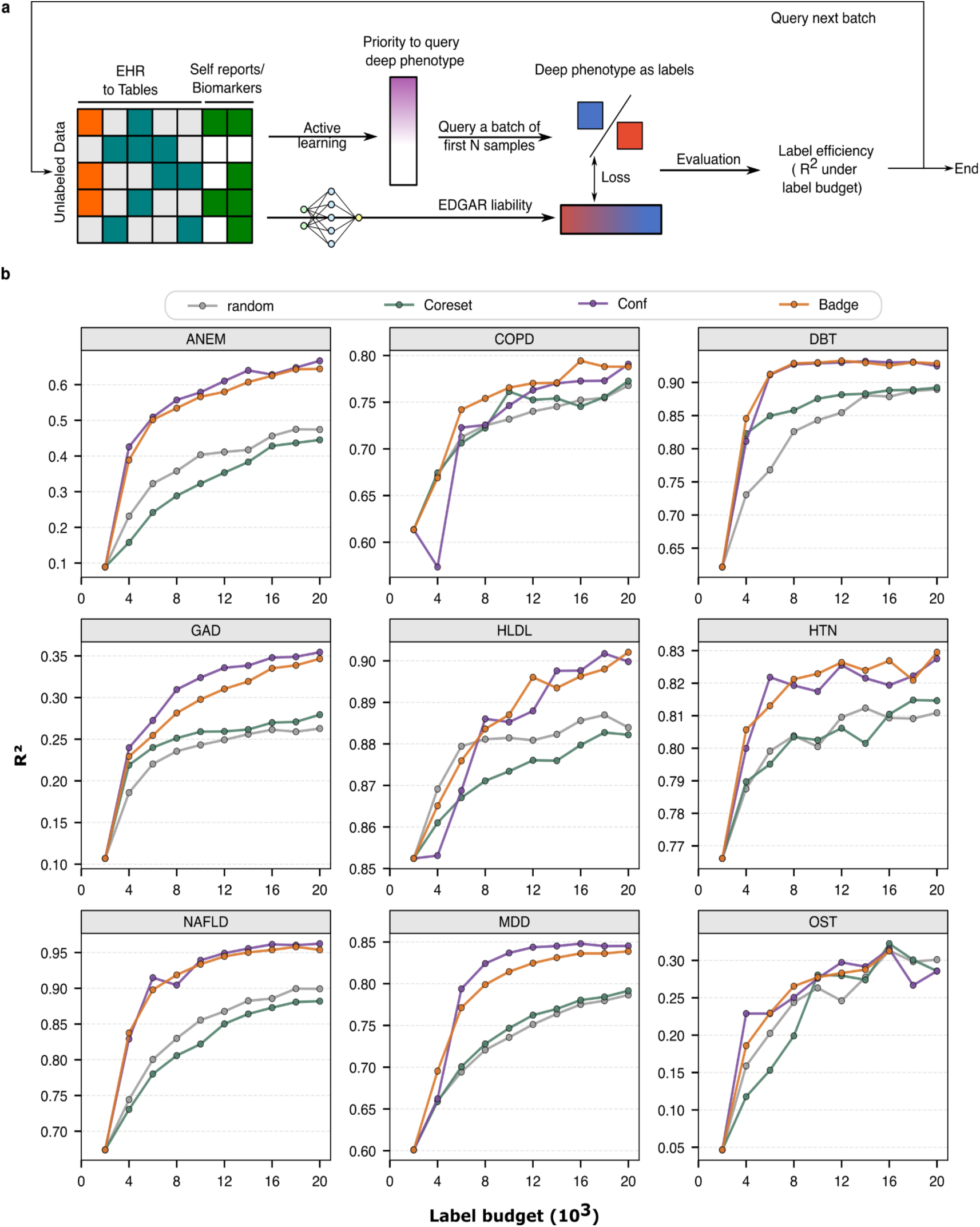
Efficient disease liability prediction enabled by deep batch active learning. **a)** Framework to use deep batch active learning to reduce the sampling burden for obtaining deep phenotype labels and thus improving label efficiency. **b)** Simulation results on UKBiobank Data. The x-axis indicates a given budget for how many samples are allowed to have labels. The y-axis shows the prediction R^2^ on the test dataset of models trained with labels selected by different active learning methods.

## Improving label efficiency with active learning in EDGAR

Intuitively, under a limited call-back budget, we should select to call back a subset of individuals that are most representative or most informative for training the EDGAR prediction model. As such, we ask if we may prioritize patients for call-back using active learning strategies, which are machine learning algorithms that identify and request the most informative samples for labeling, significantly reducing the amount of training data required to achieve high accuracy. While the use of active learning for EHR code refinement for individual diseases has been previously discussed^66^, its integration with semi-supervised learning has not been demonstrated, and its utility across diseases not previously evaluated.

We assess three different active learning approaches: Conf^67^, Coreset^68^, and Badge^69^ (**Methods,** detailed explanations of each approach in **Supplementary Methods**), and compare them to random sampling in terms of label efficiency in EDGAR predictions. Specifically, we employ each active learning strategy to EHR and disease-relevant features we use as inputs for our prediction model, and ask which individuals should be prioritized for deep phenotype acquisition (**Figure 2a**). We then “acquire” and use the deep phenotype labels of the top N prioritized individuals of each active learning strategy as labels for prediction model training, and assess the prediction model performances on held-out test dataset (**Figure 2a**).

We find that Badge and Conf outperform random sampling across diseases for most of the label budgets (N). Specifically, Conf consistently shows superior or similar R^2^ compared to other active learning methods. To achieve the same R^2^ obtained using a random sampling with a budget of N=20,000, Conf prioritizes informative individuals to label such that only 41.11% (sd=18.53%) of the 20,000 labelling budget is needed, representing a 2.43-fold improvement in label efficiency (**Figure 2b**). We find that Coreset sometimes performs worse than random sampling (for example, for ANEM, HLDL, and NAFLD; **Figure 2b**), suggesting that diversity-based active learning may not align well with semi-supervised training objectives (**Supplementary Methods**). Overall, our results show that active learning can massively reduce the budget necessary for patient call-back studies aimed at obtaining deep phenotype labels for disease liability predictions.

## EDGAR liability improves GWAS power and preserves specificity of the disease

We perform GWAS on the three phenotypes for each of the nine diseases: deep phenotypes, EHR phenotypes and our EDGAR disease liabilities (**Methods**, **Figure 3a**). Deep phenotypes produce comparatively few genome-wide significant loci for the majority of diseases, which is expected given their limited sample size and power (**Supplementary Table S2**). EHR phenotypes, defined across the full cohort, yield a higher number of significant loci than deep phenotypes for four of the nine diseases, indicating that broad phenotype availability provides moderate gains when phenotype resolution is suboptimal. EDGAR liabilities show the greatest power for GWAS locus discovery. For seven of the nine diseases, EDGAR liabilities identify more genome-wide significant loci (P < 5 × 10⁻⁸) than the matched EHR phenotypes (**Figure 3b**, **Supplementary Table S5, Supplementary Figures S10-18, Supplementary Methods**).

**Figure 3.**
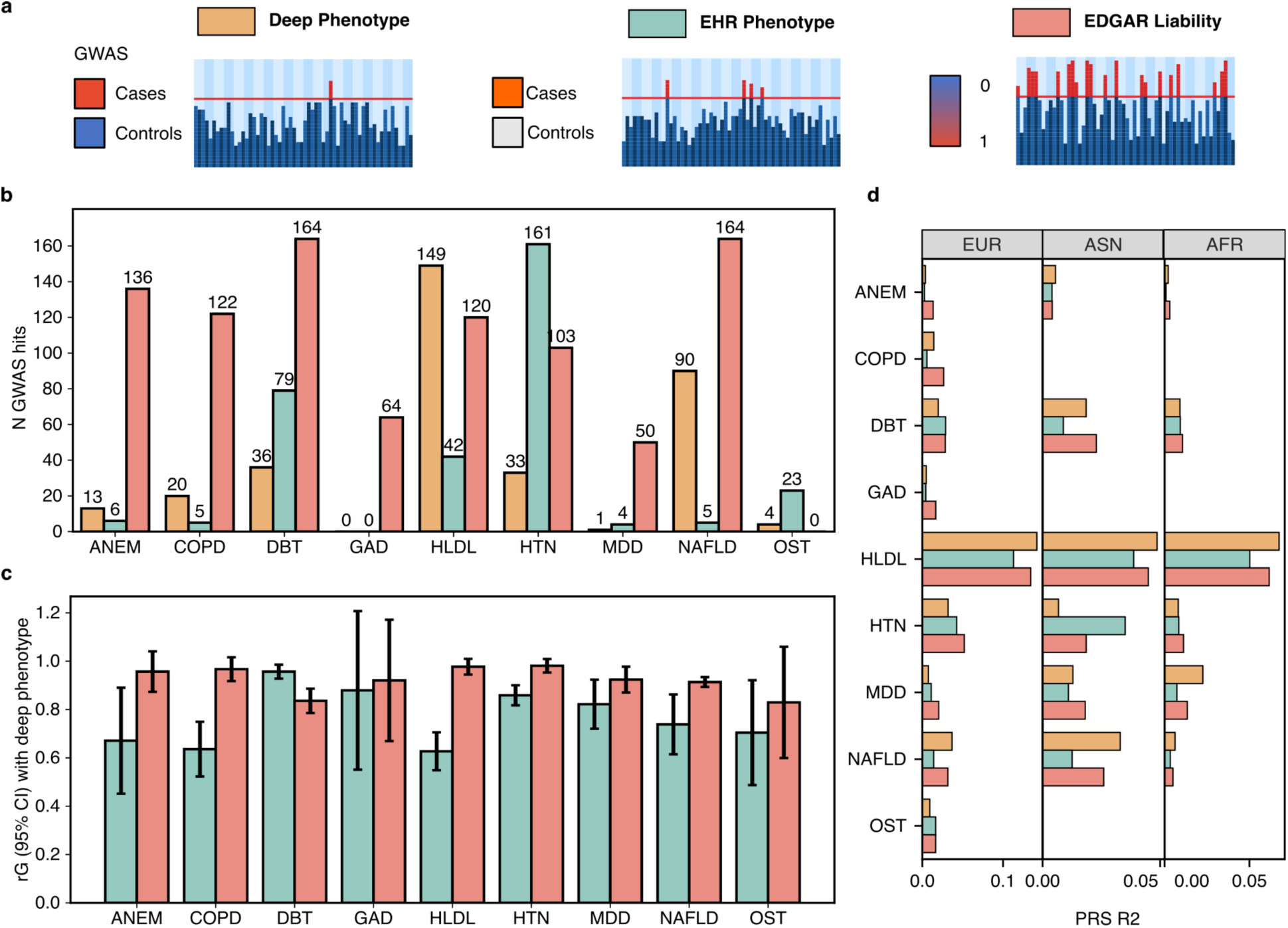
EDGAR disease liabilities increase GWAS power and preserve polygenic architecture of the deep phenotypes. **a)** Schematic of GWAS conducted on White British Europeans in UKBiobank with different phenotype definitions. Manhattan and QQ plots of all GWAS (9 diseases **⨉** 3 phenotypes) performed are in **Supplementary Figures S10-18. b)** The number of genome-wide significant hits (P<5e-8) for GWAS performed on the three phenotypes for each disease. The numbers of novel loci identified in disease liability GWAS, not found in deep phenotype GWAS, are listed in **Supplementary Table S5**. **c)** Genetic correlations between EHR and disease liability phenotypes with the deep phenotypes. **d)** Variance explained R^2^ in deep phenotypes of non-white British samples (Europeans, Asian, and Africa) using PRS derived from GWAS conducted on white British Europeans with different phenotype definitions. The sample sizes in ASN and AFR for COPD, GAD, and OST are too low (**Supplementary Table S6**) and excluded in the analyses.

To assess if the genetic effects identified via GWAS are disease relevant and specific, we perform the following analyses. First, we ask whether the EDGAR disease liabilities or EHR phenotypes have higher SNP-based genetic correlation (rG, **Methods**) with the deep phenotypes of the corresponding diseases. In eight of nine diseases, the former have higher rGs with the deep phenotype than the latter (**Figure 3c**). The only exception is DBT, which is consistent with their high phenotypic correlation between deep and EHR phenotype. This demonstrates that on the genome-wide level, genetic effects obtained with predicted disease liabilities are more consistent with those obtained using deep phenotype labels than EHR codes. Second, we ask if the polygenic risk scores (PRS, **Methods**) obtained from EDGAR disease liabilities do well at predicting the corresponding deep phenotypes out-of-sample. We first perform this prediction in the held-out set of non-White British Europeans in UKBiobank (**Supplementary Table S6, Supplementary Methods**). We find that in seven out of nine diseases, the PRS made using GWAS on EDGAR disease liabilities do better at predicting disease than those obtained from GWAS on EHR phenotypes (**Figure 3d**). We then perform the same analysis in individuals of Asian (ASN) and African (AFR) ancestries for six out of nine diseases with more than 30 cases (**Supplementary Table S6, Supplementary Methods**), asking if EDGAR predictions do better in terms of cross-ancestry PRS portability than EHR phenotype. As shown in **Figure 3d**, PRS based on EDGAR disease liabilities outperform EHR-based PRS in 11 out of 12 disease-ancestry results.

Finally, we ask how EDGAR disease liabilities compare with EHR and deep phenotypes in terms of PRS disease specificity. We obtain each of their PRS Pleiotropy^61^, the ratio of PRS R^2^ between non-disease phenotypes and target disease, using the deep disease phenotypes in UKBiobank as target phenotypes, and 147 other non-disease phenotypes as comparisons (**Methods, Supplementary Table S4**). EHR GWAS show the highest PRS Pleiotropy in seven out of nine diseases (**Extended Data Figure 2**). For seven diseases, EDGAR disease liabilities show similar or even lower PRS Pleiotropy than the deep phenotypes. We note that the exceptions (OST and GAD) with relatively higher non-specificity in EDGAR disease liabilities are the two diseases with lowest prediction quality (Pearson’s r < 0.6, **Supplementary Methods**). Overall, our results demonstrate that GWAS performed on well-modeled EDGAR disease liabilities captures disease-specific genetic effects with higher power and specificity than EHR phenotypes.

## Distinction between EHR and deep phenotype is generalizable

We next assess whether our findings generalize to external datasets. We collect two sets of GWAS results for the nine diseases for our analyses (**Figure 4a, Supplementary Methods**): EHR-based GWAS from FinnGen^70^ (External EHR GWAS, **Supplementary Table S7**), and GWAS performed on non-UKBiobank cohorts that are based on deep phenotypes (clinical assessments or biological measurements, External Deep GWAS, **Supplementary Table S8**).

**Figure 4.**
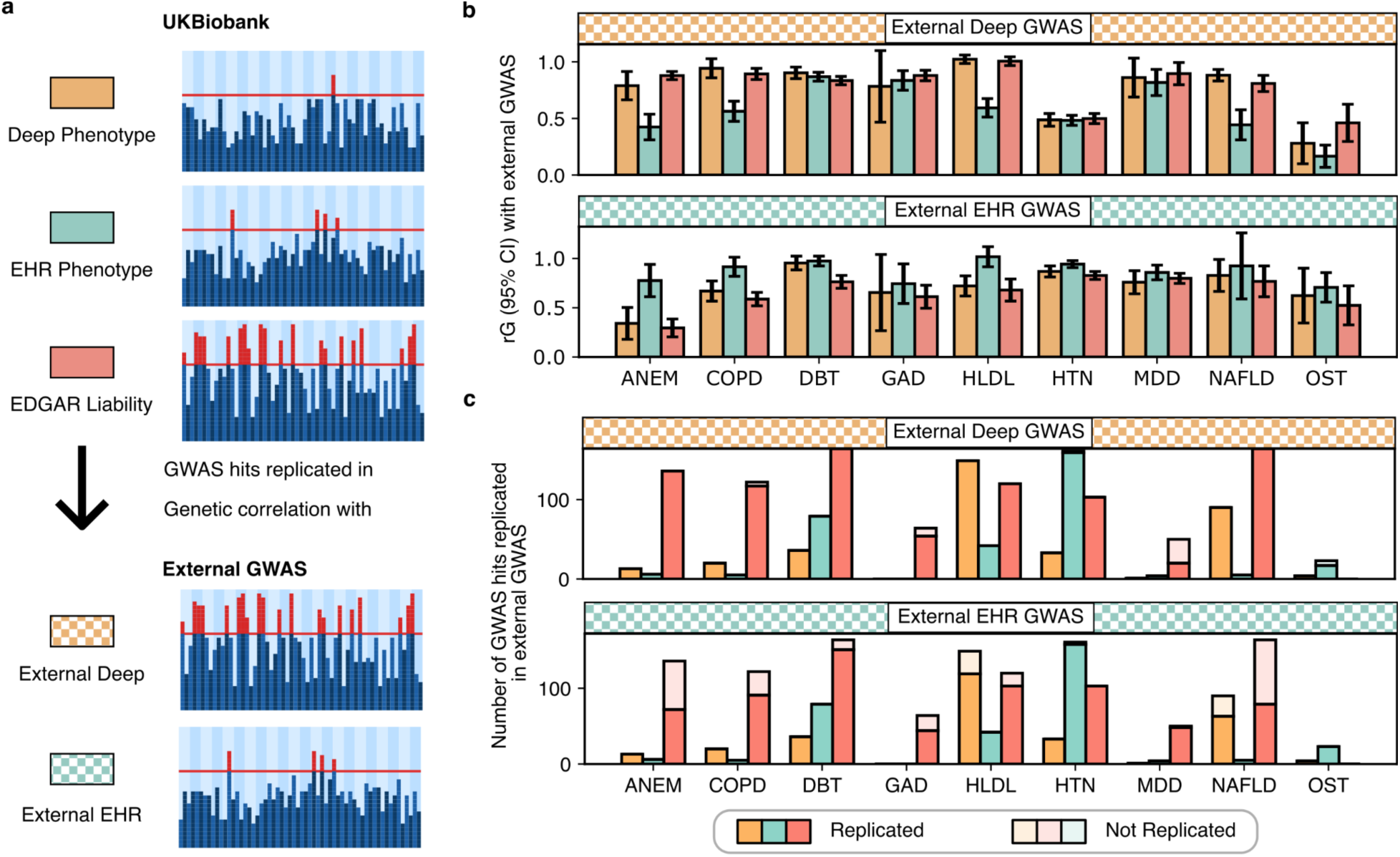
Validation in external GWAS. **a)** Schematic of GWAS results defined by data source (UKBiobank vs. External) and phenotype definition (Deep, EHR, and EDGAR liability). For each disease, we compare three UKBiobank GWAS phenotypes against two External GWAS phenotypes by (**b**) rG and (**c**) replication of GWAS hits. Replication is defined as p < 0.05 / (total number of GWAS hits from the corresponding UKBiobank GWAS). **b)** rGs between UKBiobank GWAS phenotypes and corresponding External GWAS phenotypes. **c)** Number of genome-wide significant hits identified in the internal GWAS that are replicated in the corresponding external GWAS. Replication rate (%) is in **Supplementary Figure S19.**

We first examine rGs between our three phenotypes in UKBiobank and the External Deep GWAS. While heterogeneity in phenotype definitions between cohorts may contribute to disease-specific variation, the overall pattern consistently distinguishes deep and EHR phenotypes across cohorts: in eight out of nine diseases, UKBiobank deep phenotypes show higher rGs with External Deep GWAS than UKBiobank EHR phenotypes (**Figure 4b**); in contrast, UKBiobank EHR phenotypes for all nine diseases exhibit the highest rGs with External EHR GWAS (**Figure 4b**). EDGAR disease liabilities closely track UKBiobank deep phenotypes in terms of their rGs with both External Deep GWAS and External EHR GWAS. Altogether, these results indicate that EHR phenotypes display highly replicable genetic architectures across EHR systems that exceeds their genetic sharing with deep phenotypes, likely pointing to heritable confounders^2^ that persist across EHR systems. In other words, the apparent robustness of EHR-based genetic associations may be an artifact of replicable bias.

We next examine how GWAS hits identified in our three phenotypes in UKBiobank replicate in External Deep GWAS and External EHR GWAS. When using External Deep GWAS as the replication dataset, the vast majority of genome-wide significant hits identified in all three UKBiobank phenotypes were replicated, demonstrating, in particular, the robustness of the GWAS hits we obtain from EDGAR disease liabilities (**Figure 4c**). In contrast, when using External EHR GWAS as the replication dataset, replication rates are markedly reduced for EDGAR liabilities in six out of nine diseases, including ANEM, COPD, DBT, GAD, HLDL, and NAFLD (**Figure 4c, Supplementary Figure S19**). This suggests External EHR GWAS cannot capture disease-relevant GWAS hits obtained using EDGAR GWAS, while External Deep GWAS can. Overall, replication patterns are consistent with the rG findings, further supporting a genetic difference between deep and EHR definitions that generalizes across cohorts.

## A common confounder inflates cross-disease genetic correlations in EHR GWAS

We then investigate whether there is a systemic bias, due to unrecognized heritable confounders^2^, that drives the genetic differences between EHR and deep phenotype across diseases. To test this, we compare the cross-disease rGs in EHR phenotypes against deep phenotypes within the UKBiobank. Strikingly, EHR phenotypes show inflated rG estimates as compared to deep phenotypes (**Figure 5a**, rG range -0.40 to 0.88), and are nearly all positive (**Figure 5b**, rG range -0.04 to 0.77). In contrast, rGs between EDGAR disease liabilities closely resemble those obtained from deep phenotypes (**Figure 5c**, rG range -0.47 to 0.95). While our EDGAR model does not inadvertently introduce spurious cross-disease genetic effects that inflate rGs between diseases, we hypothesize an EHR-systemic bias does^2^. This bias, if remained un-identified, would be confounded with the shared pathway between diseases.

**Figure 5.**
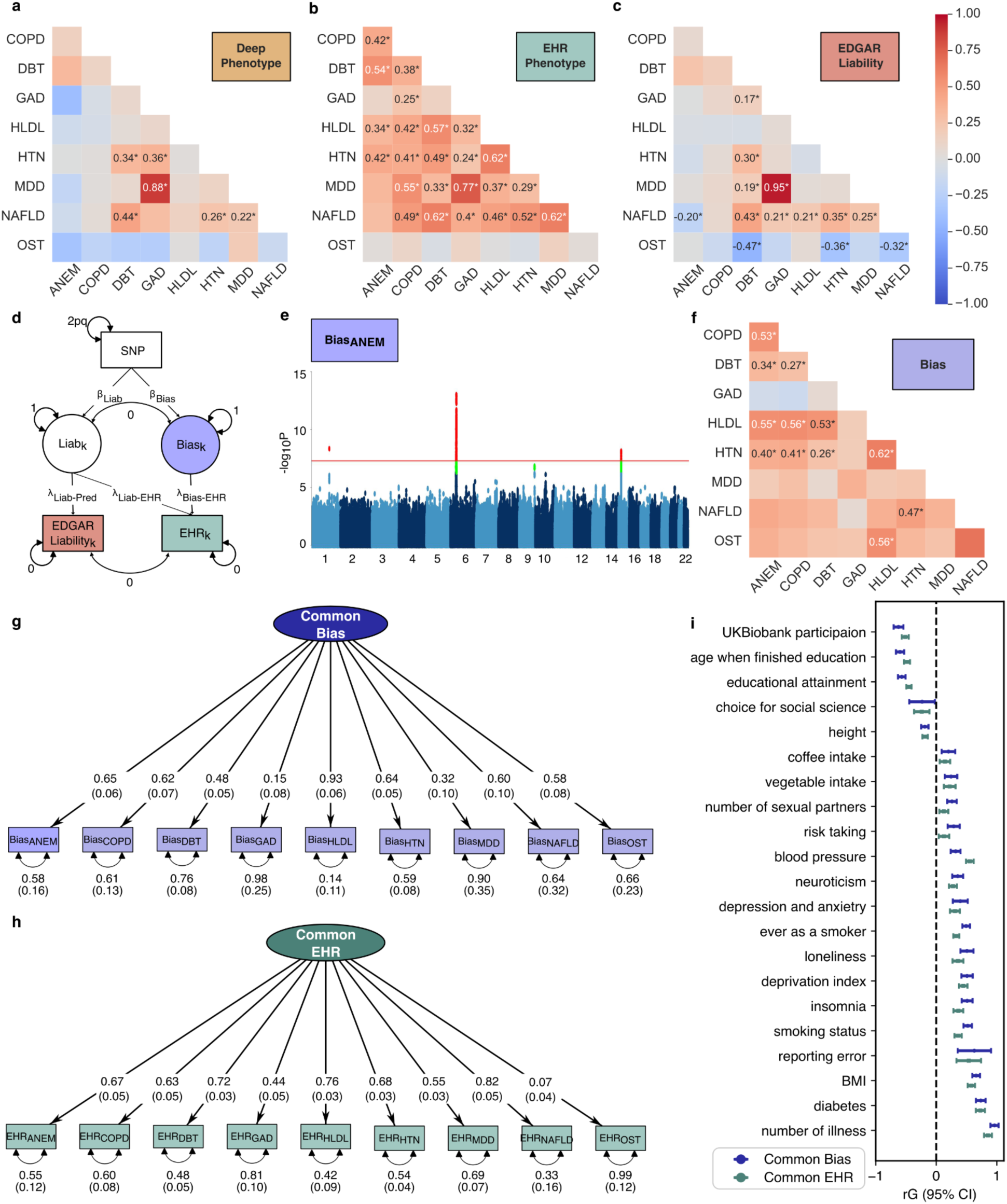
Identification of a common confounder biasing EHR rG estimates. Asterisks (*) in rG heatmaps indicate significant genetic correlations after Bonferroni correction for 144 pairwise disease rG tests. **a-c)** Cross-disease genetic correlations in Deep phenotypes (**a**), EHR phenotypes (**b**), and predicted liability phenotypes (**c**). **d)** GWAS-by-subtraction model for isolating the latent bias from latent liability. For a specific disease k, EDGAR Liability_k_ and EHR_k_ are the EDGAR disease liability and EHR phenotype of the disease, respectively. Liab_k_ is the latent disease liability that influences EDGAR Liability_k_ and EHR_k_. The Bias_k_ is the latent bias component that contaminates the EHR_k_ GWAS. **e)** Manhattan plot for Bias_ANEM_. Manhattan plots for other disease specific biases are in **Supplementary Figures S20-27**. **f)** Cross-disease genetic correlations among the Bias phenotypes extracted from (d). **g)** Standardized results from genomic SEM constructing a common latent factor (Common Bias) across disease biases. **h)** Standardized results from genomic SEM constructing a common latent factor (Common EHR) across disease EHR phenotypes. **i)** Genetic correlations of Common Bias and Common EHR with socioeconomic and behavioral traits. Full results for all tested traits in **Supplementary Table S10.**

We therefore ask whether we can identify the genetic component of this bias that contaminates the EHR GWAS. We first use the GWAS-by-subtraction^71^ framework to isolate the bias in EHR phenotype for each specific disease (**Methods**), assumed to be orthogonal to the true disease liability captured by the EDGAR disease liability (**Figure 5d**). Having derived the nine disease-specific bias factors (**Figure 5e**, **Supplementary Figure S20-27, Supplementary Table S9**), we find positive rGs among them (**Figure 5f**) that closely mirrors the rG pattern we observe among the EHR phenotypes (**Figure 5b)**.

We then fit a common factor model (Common Bias model) in genomicSEM to extract the shared heritable confounder in EHR phenotypes between diseases (**Figure 5g**, 𝜒^!^=33.05, df=27, p=0.20, AIC=69.05, CFI=0.99, SRMR=0.10). To understand what may constitute this heritable confounder, we estimate its rGs with a series of socioeconomic and behavioral traits^72–77^, many of which have previously been identified to be related to participation biases in the UKBiobank^74,75^ (**Figure 5i, Supplementary Table S10, Supplementary Methods**). We find that the Common Bias factor has significant rGs with a number of these traits, notably with worse general health (number of illness), lower educational attainment, smoking, higher mental distress, risk taking behavior, lower UKBiobank participation probability^74^, and increased self-reporting error^75^. This suggests, consistent with epidemiological evidence, that EHR-systemic bias is likely driven by healthcare engagement and operational biases^19,24,36^. We note that two disease-specific biases (GAD and MDD) show relatively low loadings on the Common Bias factor (λ<0.35, **Figure 5g**), suggesting they may have biases distinct from the rest of the diseases that may need to be investigated separately (**Supplementary Methods, Supplementary Figure S28,S29**).

Finally, we extract the Common EHR factor from the nine EHR phenotypes that index the shared genetics between them (**Figure 5h**, 𝜒^!^=444.69, df=27, p=3.69e-77, AIC=480.69, CFI=0.76, SRMR=0.11), and find that it has a high rG with the Common Bias factor (rG=0.97, SE=0.03, p=7.2e-169). Furthermore, the rGs between the Common EHR factor with socioeconomic and behavioral traits closely mirror those of the Common Bias factor (**Figure 5i, Supplementary Table S10**). These results support our hypothesis that the shared genetics between EHR phenotypes is driven by heritable confounders, and cautions that, when left unidentified, the heritable confounders in EHR phenotypes can be falsely interpreted as shared biological between diseases and socioeconomic/behavioural phenotypes, biasing our understanding of disease biology and limiting the utility of EHR-based GWAS.

## EHR Bias removal eliminates spurious genetic correlations and recovers disease liability

The Common Bias factors identified above are based solely on EHR phenotypes in UKBiobank. Two important questions remain: does this bias generalize to external EHR GWAS? And if so, can we remove them from existing GWAS performed on external EHR phenotypes? To answer this, we first examine the rGs between external EHR phenotypes for the same nine diseases from FinnGen (External EHR GWAS) and the Common Bias factor we identify among UKBiobank EHR phenotypes. We find that the External EHR GWAS for all nine diseases show positive rGs with Common Bias factor, out of which seven (except for GAD and OST) are significant, with rG=0.44-0.68 (**Figure 6a**, **Supplementary Table S11**). This suggests the Common Bias identified in UKBiobank generalizes across EHR. In contrast, many External Deep GWAS (**Supplementary Table S11**) show low or non-significant rG with the Common Bias factor (**Figure 6a**). The External Deep GWAS of DBT, GAD, MDD, and OST show modest rGs with the Common Bias factor (rG=0.35-0.52, **Figure 6a**) suggesting either there are biases in these External Deep GWAS (**Supplementary Methods**), or that the liability of one disease may bias that of another. For example, it is possible that having GAD or MDD drives the care-seeking patterns that manifest as bias in HLDL. If this is the case, then the observation that GAD and MDD disease-specific bias have low loadings onto the Common Bias factor would be consistent with the high rGs between the External Deep GWAS of GAD and MDD with the Common Bias factor. In other words, the disease liability of GAD and MDD are statistically inseparable from Common Bias in our current framework.

**Figure 6.**
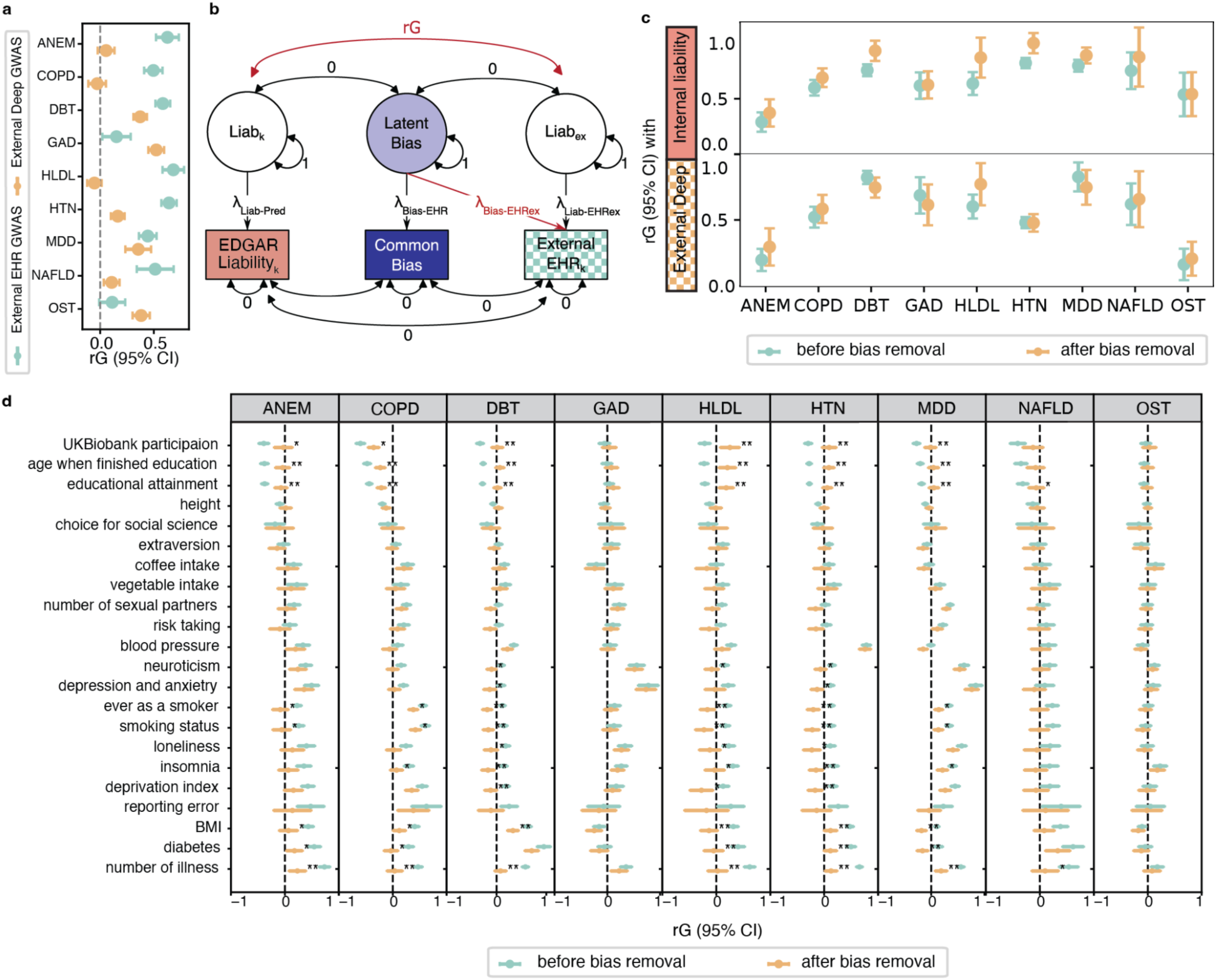
Bias removal increases genetic correlation with disease liability and eliminates spurious correlation with behavioral traits. **a)** rG between Common Bias and external deep GWAS and external EHR GWAS. **b)** GWAS-by-subtraction model for bias removal. The latent bias component is assumed to confound EHR systems beyond UK Biobank. External EHR_k_ denotes disease k phenotypes from external EHR resources (e.g., FinnGen). Liabₑₓ represents latent disease liability for External EHR_k_, independent of the latent bias. Liab_k_ represents latent disease liability measured internally from EDGAR liability. **c)** rG between external EHR with disease liability before and after bias removal. Top: disease liability inferred internally (from UK Biobank EDGAR liability); Bottom: disease liability inferred from external deep-phenotypes. **d)** Bias removal abolishes spurious rG with behavioural traits. The x-axis shows rG between external EHR and behavioural traits before and after bias removal. Single asterisk (*) denotes significant rG reduction after FDR correction (P_FDR_ < 0.05); Double asterisk (**) denotes significant rG reduction after Bonferroni correction (P_BF_ < 0.05).

Despite this caveat, we derive a bias correction model to remove the bias identified in one EHR GWAS from another. In this model, we assume that the Common Bias factor, identified in UKBiobank, is a generalizable bias that also influences External EHR GWAS (in this case from FinnGen) and that is statistically separable (orthogonal) to latent disease liabilities captured by the External EHR GWAS or EDGAR disease liabilities in UKBiobank (**Figure 6b**). With this assumption, we can easily remove the latent bias from External EHR GWAS using GWAS-by-subtraction (**Figure 6b, Methods**). We first show that removing this latent bias leads to increases in rG between External EHR phenotypes and EDGAR disease liabilities in the UKBiobank (**Figure 6c**). While this is expected (**Supplementary Methods**), we also find that the bias removal leads to increased rGs between External EHR and External Deep GWAS for six out of nine diseases (**Figure 6c**). The three exceptions (DBT, GAD, and MDD) are those whose disease liability and systematic bias may be statistically inseparable (**Figure 6a**). Finally, we find that bias removal from External EHR phenotypes significantly reduces their rGs with socioeconomic and behavioral traits, such as number of illnesses, education, and UKBB participation (**Figure 6d** and **Supplementary Table S12**).

Altogether, our results provide a proof of concept for removing bias from GWAS summary statistics derived from one EHR using systematic bias identified from another, while showcasing the limitations of this approach on diseases whose liabilities form part of the systematic bias. Notably, the resultant GWAS from our bias removal approach captures, like all results of GWAS-by-subtraction, only the fraction of genetic effects on individual diseases that are not shared with the Common Bias factor. It no longer captures all genetic effects on the individual diseases.

## Discussion

Despite a large literature on biases in healthcare data^14,15,24–27^, the use of EHR codes in GWAS ^1,8–10^ and deep-learning models for disease predictions^50–53^ have become more prevalent with the increasing availability of EHR data for research use. In this paper, we propose and demonstrate the effectiveness of EDGAR, a disease liability prediction model that integrates clinically-assessed and disease-relevant data, and that can be augmented by active learning for training label selection. We demonstrate that EDGAR can improve the phenotypes we derive from EHR for genetic analyses: GWAS on EDGAR disease liabilities outperform GWAS on both deep and EHR phenotypes in terms of power, out-of-sample PRS prediction accuracy and PRS prediction specificity. We further showcase how they may be used in factor models to identify systemic biases in EHR phenotypes, thereby facilitating bias-removal from any existing EHR-based GWAS.

Notably, our model aims to learn lifetime disease liability, which works specifically in service for genetic studies, not for prediction of future EHR events. We note that there are previous works on disease phenotype imputation^60,61^, refinement^54^ and liability prediction^78^ for genetic studies. However, those that can clearly demonstrate the imputed or predicted disease liabilities capture more disease-specific genetics have only been shown to work through leveraging rich biobank phenotypes^60,61^ or disease-specific measures^54^ (as opposed to all available EHR events). Conversely, those that work on EHR events to predict masked or next EHR events have not aimed to predict disease liabilities that would be suitable for identifying disease-specific genetics, and they tend to propagate biases in EHR as they do not make use of deep phenotypes labels for model training^53,78^. Our model avoids the otherwise inevitable consequence of models trained only on EHR codes propagating EHR-systemic biases into its predictions. Further, our use of active learning methods to prioritize individuals for obtaining deep phenotype labels can be instructive for future prospective recall studies that can only be performed with a fixed budget. Overall, our unified framework for disease liability prediction presents a feasible way for deriving lifetime disease liabilities from existing EHR diagnostic codes specifically for identifying disease-specific genetic associations.

While our results demonstrate that tabular architectures can perform just as well as attention-based sequential models for sparse EHR events, the growing availability and resolution of longitudinal data beyond diagnosis (e.g. drug prescriptions and key metabolites changing in levels due to treatments) presents further opportunities to model the underlying continuous nature of diseases progression. Future work may focus on exploring continuous-time frameworks^79–81^ that can more flexibly capture the stochastic evolution of these diverse clinical trajectories and the modulatory relationships between diseases.

In terms of genetic findings, EDGAR disease liabilities show greater numbers of replicable GWAS hits (in External Deep Phenotypes), better out-of-sample and cross-ancestry PRS prediction, and higher PRS prediction specificity to target disease than EHR phenotypes. Beyond what this all means for GWAS discovery, this enables us to isolate method-specific bias from underlying disease-specific etiology: the identification of a systemic, heritable confounder across diseases that induces high rGs between EHR phenotypes. This systemic bias has high rGs with a series of traits reflecting healthcare-seeking behavior, reporting patterns, and socioeconomic context. We note that previous studies^74,75,82–84^ have made attempts to mitigate known confounding sources like misdiagnosis between different diseases. By contrast, our approach does not require prior knowledge of the likely sources of confounding factors or the extent to which they bias GWAS findings. Rather, it leverages two independent assessment modalities (observed EHR phenotypes and EDGAR disease liabilities aligned to deep phenotypes) to statistically decompose the observed EHR phenotype into latent disease-specific liability, which is informative of disease-specific genetics, and method-specific bias^13^, which is not. As our approach is dependent on the availability of good (disease-specific) disease liability phenotypes from which EHR-based biases may be isolated, it will be further improved as future liability predictions produce better approximations of true disease liability. In particular, we note that our curated deep phenotype disease labels, used for model training, are constrained by data availability and subject to participation bias just like the rest of the data in UKBiobank (though potentially different from those specific to EHR events), and should not be regarded as universally valid ground truth (**Supplementary Methods**). We further discuss the consequences of imperfect disease liability prediction for our bias identification model in **Supplementary Methods**.

Crucially, our results indicate that systemic biases in EHR-derived phenotypes are, to a substantial extent, shared across EHRs and can be mitigated using summary-level genetic association statistics alone. Removing the systemic bias factor learned in UKBiobank from FinnGen EHR GWAS can attenuate the latter’s spurious associations with socioeconomic traits, providing a proof of concept consistent with theoretical expectations. Future work may explore more robust Genomic SEM specifications informed by known disease relationships (e.g., psychiatric-specific bias; **Supplementary Methods**) to better decouple latent confounding architecture from genuine biological pleiotropy. Further, we note that both EHRs we investigate, UKBiobank and FinnGen, are from European countries. While the systemic bias factor learned in UKBiobank may help attenuate biases in FinnGen, it remains to be determined in future investigations whether EHRs in other populations may be similarly amenable.

Overall, our EDGAR framework predicts lifetime disease liabilities that are disease-relevant and improves GWAS and PRS predictions, and enables the identification and correction of heritable confounders in EHRs data and alignment with gold-standard disease definitions. These findings have broad implications for large-scale GWAS: whenever EHR-based case definitions are used, systematic confounding should be assessed and corrected to avoid misleading biological conclusions.

## Methods

### Ethical approval

This research was conducted under the ethical approval from the UKBiobank under applications 28709 and 163937.

### Samples and phenotypes

This study uses data from 337,121 white British Europeans in the UKBiobank^85^. We focus on nine diseases, including two psychiatric disorders, generalized anxiety disorder (GAD) and major depressive disorder (MDD), as well as seven non-psychiatric diseases: anaemia (ANEM), chronic obstructive pulmonary disease (COPD), hyper-LDL-cholesterolemia (HLDL), non-alcoholic fatty liver disease (NAFLD), diabetes (DBT, inclusive of Type 1 and Type 2), hypertension (HTN), and osteoporosis (OST). We define the EHR phenotype for each disease as the presence of at least one diagnosis code that maps to the corresponding ICD10 definition. We extract all diagnosis codes from both primary care (GP) in Read2 and CTV3 formats and inpatient records (HES) in ICD9 and ICD10 codes (**Figure 1a**). The prevalences of each disease, across all diagnostic codes available, are summarised in **Supplementary Table S1**. Beyond diagnoses, we identify additional disease-relevant features, summarized in **Supplementary Table S1** and **Supplementary Table S4**. Briefly, these included: blood cell count and biochemistry measures for ANEM, HLDL, NAFLD, and DBT; spirometry for COPD; blood pressure for HTN; bone mineral density (BMD) from DEXA scans for OST; and questionnaire-based mental health and lifestyle variables for GAD and MDD. Finally, we derive deep phenotypes for each of the nine diseases according to diagnostic criteria obtained from medical literature or previous GWAS performed on clinician assessed cohorts^16,86–95^ (**Supplementary Table S2)**.

### The EDGAR framework

In EDGAR, we adapted the AutoComplete^60^ architecture by shifting its objective function from broad-spectrum data imputation to target-specific loss optimization. Whereas the original model employs a self-supervised approach to reconstruct all missing entries across the input matrix (encompassing all features and labels), our adaptation constrains the loss function to be supervised exclusively by masked deep phenotype labels and additional clinical features (when included) to ensure the model is not affected by noisy EHR features.

To enable this model to work on sequential EHR diagnostic codes, we convert the latter into counts of each diagnostic code per individual, which can be represented in a tabular format. Specifically, we mapped ICD-9 codes into ICD-10 because they are sparse. In contrast, we did not merge Read2 and CTV3 codes, as they are sufficiently dense and their original format carries operational information that may be informative for prediction (e.g., version of the health system, event context such as GP vs. HES). As a result, there are 301,352 individuals with at least one of the 1,602 ICD-10 codes from HES, 45,929 had at least one of the 1,987 GP Read 2 codes, and 112,255 had at least one of the 3,974 GP CTV3 codes. When converting these EHR codes into tabular counts, temporal information was lost. To partially preserve it, we added the age at first and age at last occurrence for each code. Specifically, 1,602, 1,987, and 3,974 columns representing age at first occurrence were added for HES ICD-10, GP Read2, and GP CTV3 codes, respectively. Because many codes occur only once for a given individual, age at last occurrence was included only for high-frequency codes, defined as codes that occur more than once in at least 1% of individuals. As a result, 143, 856, and 781 columns of age at last occurrence were added for HES ICD-10, GP Read2, and GP CTV3 codes, respectively.

We train the model on 90% of the samples and evaluate on the rest 10%. We use Pearson’s r to evaluate predictiveness of each disease liability prediction for deep phenotype labels, and macro-AUC (the average AUC across nine diseases) to provide an overall predictiveness evaluation of the model’s ability compared to other studies^53^. We use the same parameters for AutoComplete training as in the previous work (**Supplementary Notes**); model training is conducted on a single NVIDIA RTX 4090 GPU. We compute SHAP^64^ values post-hoc to quantify the contribution of input features to the disease liability predictions (**Supplementary Figure S1-9**).

### Active learning

We conduct deep batch active learning experiments (**Figure 2a**) with our semi-supervised EDGAR framework. At initialization (round 0), the label column is entirely missing. To warm-start, we reveal a batch of B = 2,000 random labels (round 1). In each subsequent round, we select an additional batch of B = 2,000 unlabeled samples from the pool, “recalling” these patients to obtain deep phenotype labels, retraining the model on the full cohort with the expanded labeled subset, and evaluating the predictions on a fixed 10% test split (using variance explained R^2^). This acquisition process is repeated for 10 rounds under different strategies: a) Coreset^68^, b) Conf^67^, and c) Badge^69^ (detailed in **Supplementary Methods**). We compare the disease liability predictions with those obtained using random sampling with the same budget. All active learning experiments are conducted on a single NVIDIA RTX 4090 GPU.

#### GWAS

We perform GWAS and downstream genetic analyses on the observed EHR/deep phenotypes and EDGAR disease liabilities aggregated from ten-fold cross validation. GWAS are performed using imputed genotype data at 5,781,354 SNPs (minor allele frequency > 5%, INFO score > 0.9) using logistic regression and linear regression implemented in PLINK v2^96^ for binary and quantitative traits (predicted disease liabilities), respectively. We use 20 PCs computed with flashPCA^97^ on 337,129 unrelated White-British Europeans in UKBiobank and genotyping arrays as covariates for all GWASs (see **Supplementary Notes** for details of sample and genotype quality control in UKBiobank).

### SNP-based heritability and genetic correlation

To test for SNP-based heritability of each phenotype and the genetic correlation (rG) between pairs of phenotypes, linkage disequilibrium (LD) score regression implemented in LDSC v1.0.11^98^ was performed on the GWAS summary statistics using in-sample LD scores estimated in 10,000 random white British UKBiobank individuals at SNPs with minor allele frequency > 5% as reference. In estimating liability-scale heritability for each disease, we assume the population prevalence of binary phenotypes equaled their prevalence in UKBiobank (**Supplementary Table S1**).

### Polygenic risk scores (PRS) and PRS Pleiotropy

We use PRSice v2 (ref^99^) to calculate the PRS for non-white British samples (including Europeans, Asian, and Africa). We drive PRS from GWAS performed on unrelated White-British Europeans, thresholded at multiple P-values (from 5e-8 to 1, with an interval of 5e-5). For all PRS predictions, we used 20 genomic PCs as covariates, and evaluated accuracy in predicting the deep phenotypes using Nagelkerke’s R^2^. The final reported PRS corresponds to the best-performing P value threshold. Due to limited sample size (**Supplementary Table 6**), we perform PRS predictions for only six out of nine diseases (case N for deep phenotype >30) in Asian and African groups.

PRS pleiotropy is a specificity metric defined as the ratio of variance explained in the target disease relative to secondary phenotypes^61^. We perform ten-fold cross validation for PRS pleiotropy of the three phenotypes (EHR/Deep/EDGAR) across nine diseases. Each fold we use 90% of the individuals with the corresponding phenotype and then build the PRS from the GWAS results with PRSice v2 (ref^99^) and evaluate predictive accuracy for the focal deep phenotype and other secondary phenotypes in the 10% of individuals who were held out. Secondary phenotypes include 147 phenotypes selected from the additional features (**Supplementary Table S4)** that reflect non-etiological signals. For all PRS predictions, we used 20 genomic PCs and the genotyping array used as covariates. For binary phenotypes, we evaluated accuracy using Nagelkerke’s R^2^. For all quantitative phenotypes, we evaluated accuracy using ordinary R^2^.

### Bias identification

For each disease k, the GWAS-by-subtraction model (**Figure 5d**) specifies two latent variables—*Liab_k_* (latent disease liability) and *Bias_k_* (latent bias)—that together explain variation in two observed GWAS results: EDGAR Liability_k_ and EHR_k_. EDGAR Liability_k_ represents the GWAS for the EDGAR disease liability, and EHR_k_ represents the GWAS for the corresponding EHR phenotype. In the structural model, *Liab_k_* loads onto both EDGAR Liabilityₖ and EHRₖ, capturing the shared genetic signal reflecting true disease liability. *Bias_k_*, in contrast, loads only onto EHR_k_ and represents genetic effects that contaminate the EHR phenotype but are independent of true disease liability. We then derive a Common Bias factor by fitting a common factor model in genomic SEM across all *Bias_k_*. Detailed methodological considerations and alternative model assumptions are described in **Supplementary Methods**.

### Bias removal from external EHR GWAS

We remove the Common Bias factor identified in UKBiobank data from existing FinnGen EHR GWAS on the same nine diseases (**Supplementary Table S7**). Before bias correction, we assume that the Common Bias factor does *not* generalize to the external GWAS. Accordingly, we fix the loading of *Latent Bias* onto EHRₑₓ (𝜆_"#$%&’()*+_) to zero. Under this null specification, the genetic correlation (*rG_before_*) between *Liab_ex_* and *Liab_k_* corresponds to the observed rG between EHRₑₓ and *Liab_k_*. After removal, we allow 𝜆_"#$%&’()*+_ to be freely estimated from the genetic covariance matrix, enabling the bias factor to contribute to the external GWAS. We then get an estimated rG after bias removal (*rG_after_*). We prove that *rG_after_* is always larger than *rG_before_* in **Supplementary Methods**. We also assess whether bias removal can enhance rG with External Deep GWAS and eliminate spurious correlations with socioeconomic/behavior traits (**Supplementary Figure S30**). Z difference tests are performed to test for significance in difference between rG with socioeconomic/behavioral traits before and after bias removal; P values are Bonferroni-corrected for 387 tests (9 disease × 43 socioeconomic/behavioral traits).

## Author Contributions

Y.D. implemented the methods. Y.D. and N.C. analyzed the data. Y.D and N.C. interpreted the results.

Y.D. and N.C. wrote the manuscript. N.C. conceived the study and supervised the work.

## Data Availability

UK Biobank genotype and phenotype data used in this study are from the full release (imputation version 3) of the UKBiobank Resource obtained under application no. 28709 and 163937. We used publicly available summary statistics from FinnGen (https://www.finngen.fi/en/access_results) and other studies (downloaded from the GWAS Catalog, https://www.ebi.ac.uk/gwas/), with references in Supplementary Table S7-8. Summary statistics of all GWAS we have performed in this paper are available at https://doi.org/10.5281/zenodo.18402904.

https://www.ebi.ac.uk/gwas/

https://www.finngen.fi/en/access_results

## Acknowledgements

The authors gratefully thank all participants of UKBiobank and FinnGen who made this work possible. The authors thank Vassilis Skaramagkas, Ulzee An and Juan De la Hoz for preliminary work that facilitated this study, as well as Andy Dahl, Jonathan Flint, Kenneth Kendler, Tian Ge and Sriram Sankararaman for helpful discussions.

## Use of artificial intelligence

The authors used the large language model GPT-4 (https://chat.openai.com/) and Gemini-3 (https://gemini.google.com/) for language improvements in the preparation of the manuscript. Language models were not used to generate contributions to the original research, data analysis, or interpretation of results. All final content decisions and responsibilities rest with the authors.

## Ethical approval

This research was conducted under the ethical approval from the UK Biobank Resource under application no. 28709 and 163937.

## Data availability

UK Biobank genotype and phenotype data used in this study are from the full release (imputation version 3) of the UKBiobank Resource obtained under application no. 28709 and 163937. We used publicly available summary statistics from FinnGen (https://www.finngen.fi/en/access_results) and other studies (downloaded from the GWAS Catalog, https://www.ebi.ac.uk/gwas/), with references in **Supplementary Table S7-8**. Summary statistics of all GWAS we have performed in this paper are available at https://doi.org/10.5281/zenodo.18402904.

## Code availability

Publicly available tools that are used in data analyses are described wherever relevant in Methods and Reporting Summary. The EDGAR model is available at: https://github.com/yazhengdi/EDGAR; our benchmarking models as well as auxiliary codes are available at https://doi.org/10.5281/zenodo.18402904; the AutoComplete model we adapted EDGAR from is available at https://github.com/sriramlab/AutoComplete as previously published; the BADGE active learning implementation is available at https://github.com/JordanAsh/badge.

---

**Extended Data Figure 1.**
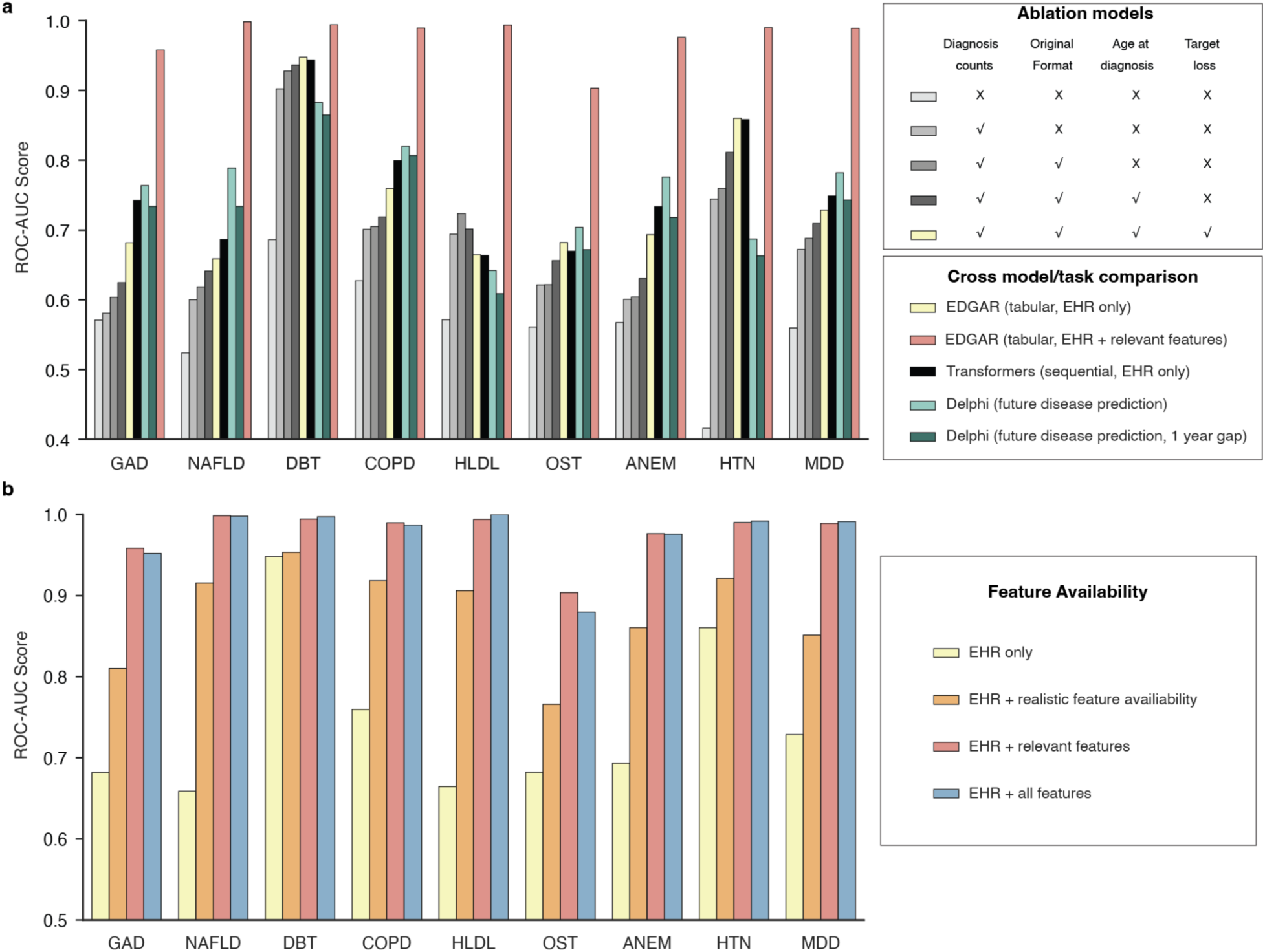
Evaluation of model performance across model architecture, task and data input. **a)** Disease-specific AUCs across our EDGAR model, in-house transformer model and Delphi. **b)** Disease-specific AUCs of EDGAR under various scenarios of feature availability. Realistic feature availability: feature availability is constrained such that additional features are only observed for patients with prior diagnostic history, resulting in a sample retention rate of 52.8%.

**Extended Data Figure 2.**
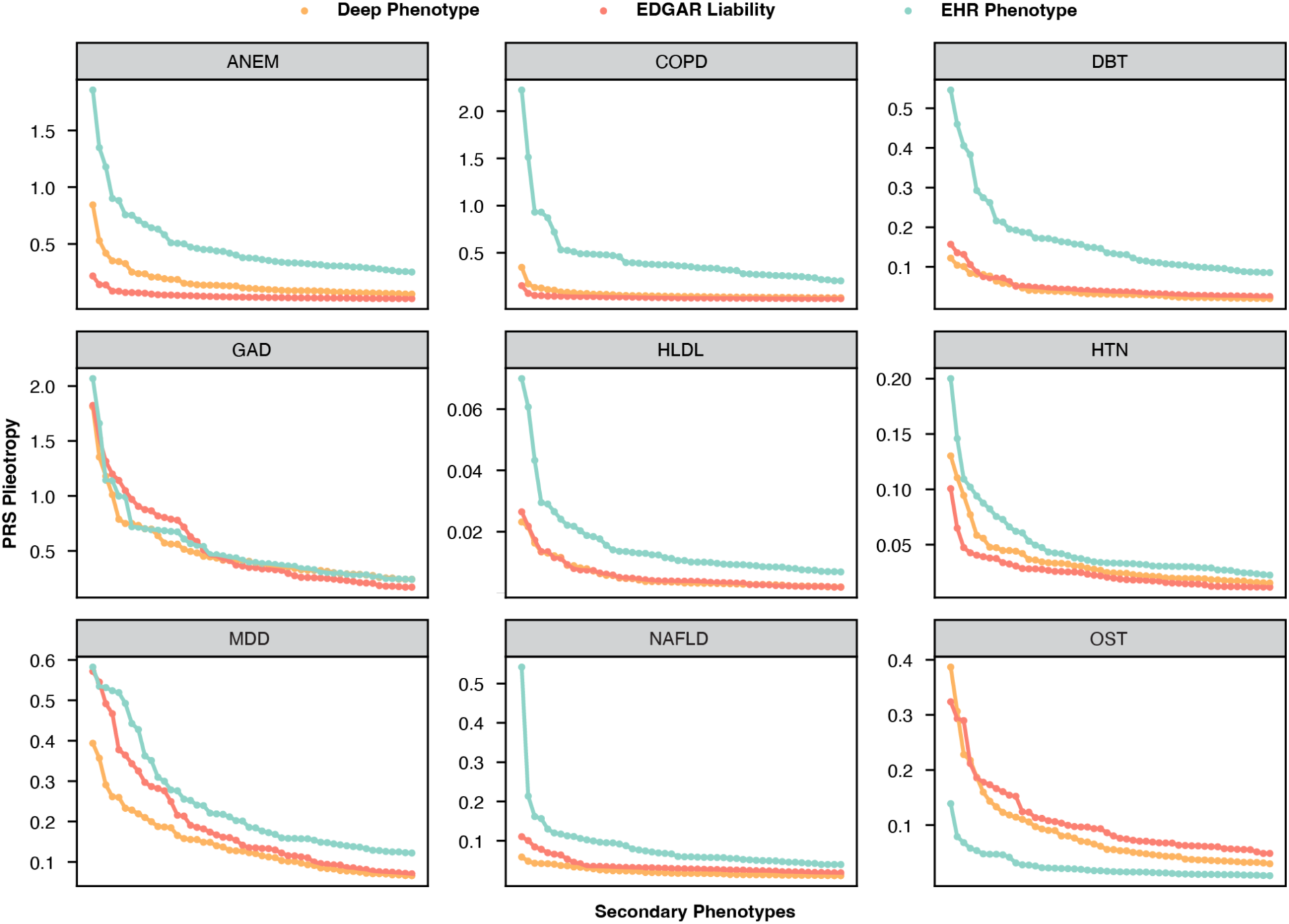
Phenome-wide PRS pleiotropy quantifies nonspecificity across phenotypes. PRS pleiotropy is defined as the ratio of PRS prediction accuracy for secondary phenotypes relative to the focal disease (PRS pleiotropy = R^2^_secondary_/R^2^_disease_). We calculate PRS pleiotropy for 147 non-disease secondary phenotypes and display the top 50 in decreasing order of rank.

