## SupplementaryMaterials for "Learning lifetime disease liability reveals and removes genetic confounding in electronic health records"

Yazheng Di[1], Na Cai[1,*]

1. Department of Biosystems Science and Engineering, ETH Zurich

*Corresponding author

**Supplementary Materials**

### Supplementary Methods

#### UKBiobank

##### *Sample filtering*

Of all 502,637 samples in UKBiobank[1] full release, we performed the following QC steps to select the samples for use in our analyses. We first removed samples that were not included in the UKBiobank full release PCA analysis, which includes samples that were indicated as "het.missing.outliers" ("Indicates samples identified as outliers in heterozygosity and missing rates, which indicates poor-quality genotypes for these samples"), "excess.relatives" ("Indicates samples which have more than 10 putative third-degree relatives in the kinship table"), and whose "Submitted.Gender" were different from "Inferred.Gender". Applying these filters brought the sample size down to 407,219. We checked that the remaining sample contains only one out of any pair or group of related individuals with relatedness > 0.05. We then selected samples indicated to be "in.white.British.ancestry.subset" ("Indicates samples who self-reported 'White British' and have very similar genetic ancestry based on a principal components analysis of the genotypes"), resulting in a sample size of 337,545.

We then removed 337 samples indicated as having "putative.sex.chromosome.aneuploidy" ("Indicates samples identified as putatively carrying sex chromosome configurations that are not either XX or XY"). Finally, we removed 79 samples who have withdrawn their consent for use of their genetic data in analyses, arriving at our final set of 337,129 individuals passing QC1. Of these samples, 37,041 were part of UK Biobank Lung Exome Variant Evaluation (UKBiLEVE)[2], a study for chronic obstructive pulmonary disease (COPD). We retain all samples in UKBiLEVE, but as they are genotyped using a custom array optimised for coverage over regions implicated in lung health and disease, we consistently use 'genotyping array' as a covariate in all our analyses.

##### *Genotype quality control*

We performed stringent filtering on imputed variants (version 3) used for GWAS in this study, removing all insertions and deletions (INDELs) and multi-allelic SNPs: we hard-called genotypes from imputed dosages at 9,720,420 biallelic SNPs with imputation INFO score greater than 0.9, MAF greater than 0.1%, and P value for violation of Hardy-Weinberg equilibrium > $10^{-6}$, in individuals with a genotype probability threshold of 0.9 (individuals with genotype probabilities below 0.9 would be assigned a missing genotype). Of these, 5,776,313 SNPs are common (MAF > 5%). We consistently use these SNPs for all analyses in this study.

We performed principal component analysis (PCA) on directly genotyped SNPs from samples in UKBiobank and used PCs as covariates in all our analyses to control for population structure using flashPCA[3]. From the array genotype data, we first removed all samples who did not pass QC, leaving 337,129 White-British, unrelated samples. We then removed SNPs not included in the phasing and imputation and retained those with minor allele frequencies (MAF) >= 0.1%, and P value for violation of Hardy-Weinberg equilibrium > $10^{-6}$, leaving 593,300 SNPs. We then removed 20,567 SNPs that are in known structural variants (SVs) and the major histocompatibility complex (MHC) as recommended by UKBiobank[1], leaving 572,733 SNPs. Of these, 334,702 are common (MAF > 5%), and from these common SNPs we further filtered based on missingness <0.02 and pairwise LD r2 < 0.1 with SNPs in a sliding window of 1000 SNPs to obtain 68,619 LD-pruned SNPs for computing PCs using flashPCA. We obtained 20 PCs, their eigenvalues, loadings and variance explained, and consistently use these PCs as covariates for all our genetic analyses.

***Disease selection***

To evaluate the robustness and generalizability of our liability prediction model, we selected nine common complex diseases based on three pre-specified criteria: (i) clinical prevalence, ensuring sufficient statistical power for population-scale genetic analysis and EHR-based inference, as rare diseases with extreme data sparsity require distinct algorithmic architectures; (ii) data availability, requiring the presence of clinical measurements (laboratory tests or physiological assessments) to serve as deep phenotype labels for supervision; and (iii) pathophysiological diversity, encompassing a broad range of organ systems and diagnostic modalities. The final selection represents a diverse clinical spectrum, including psychiatric (GAD, MDD), metabolic (DBT, HLDL, NAFLD), cardiovascular (HTN), respiratory (COPD), hematologic (ANEM), and musculoskeletal (OST) domains. This diversity allows for a comprehensive assessment of whether our framework can effectively decouple disease liability from EHR-specific confounders across varying clinical contexts.

We show our prediction framework is robust through its performance across varying levels of data availability that reflect real-world clinical informatics challenges. The wide range of input EHR prevalence (from 3.27% for NAFLD to 35.41% for Hypertension, **Supplementary Table S1**) captures the actual diagnostic distributions and inherent reporting biases within health systems. Similarly, the substantial variance in the availability of additional features (**Supplementary Table S4**), such as questionnaire response rates, reflects the varied success of recall efforts across different patient populations. Furthermore, the deep phenotype labels used for supervision demonstrate significant heterogeneity in both sample size and case-control balance (**Supplementary Table S2**), ranging from high data availability, high prevalence for HLDL (N=203,786, case ratio 47.56%) to sparse data for OST (N=28,665, case ratio 8.42%), as well as high availability, low prevalence for DBT (N=309,586, case ratio 3.73%). By incorporating this range of feature and label sparsity, we aim to characterize the framework's behavior across a wide spectrum of data availability typically encountered in biobank research. Consequently, performance gradients observed in diseases, such as the lower prediction accuracy for OST, should be contextualized as an expected result of underlying data scarcity rather than a limitation of the model architecture. Overall, these patterns underscore the inherent complexity of real-world clinical data, where a fundamental trade-off exists between the cost of acquiring deep phenotypes and the resulting predictive precision—a challenge that directly motivates the implementation of our active learning strategy.

***Deep phenotypes***

Our curated deep phenotype disease labels were constrained by data availability and should not be regarded as universally valid ground truth. For instance, the deep phenotypes for GAD and MDD were derived from CIDI-adapted questionnaire items and thus remain subject to self-report biases[4] such as recall inaccuracy and participation effects. Our DBT deep phenotype by definition cannot ditingushi between type I diabetes (T1D) and type II diabetes (T2D). But due to a low prevalence of T1D in UK Biobank compared with T2D, as well as genetic correlations (FinGen T1D: rG = 0.04, SE = 0.07, p = 0.51; FinGen T2D: rG = 0.95, SE = 0.04, p = 4.4e-158), it should mostly reflect T2D liabilities. Similarly, biomarker-based disease labels may be confounded by medication use: lipid-lowering or antihypertensive drugs can normalize biomarker levels, making one look like a control even when one has an underlying disease. In our study, we adjusted for medication use, but recognize this is not failproof. Accordingly, we treat our effort to obtain deep phenotype labels from disease-relevant data as representations of true gold-standard labels and what the roles they can serve in our model, rather than an attempt to reconstruct the actual disease ground truth. As such, when applying this framework to other diseases or cohorts, researchers should instead define study-specific gold-standard phenotypes that best capture the biological constructs of interest.

***PRS prediction in non-White British individuals in UKBiobank***

We identified 150,213 individuals who did not self-identify as White British in the UKBiobank[1]. We removed from their imputed genotypes regions of the genome that contained previously identified structural variants[5], then hard-called genotypes from imputed dosages at 5,327,974 biallelic SNPs with imputation INFO score greater than 0.9, MAF greater than 0.1%, and P value for violation of Hardy-Weinberg equilibrium > 10-6, with a genotype probability threshold of 0.9 (anything below would be considered missing).

To identify participants of European (EUR), African (AFR) and East Asian (ASN) ancestries, we selected 70,718 LD-pruned common SNPs (MAF > 5%, P value for HWE > $10^{-6}$, LD r2 < 0.1) the autosomes that overlap with SNPs in the 1000 Genomes Project Phase 3 (1000G)[6] from 150,213 non-White British individuals in UKBiobank, then projected all individuals onto PCs obtained from 1000G samples using loadings at these SNPs calculated with LDAK v5[7]. We identified 122,710, 8,040 and 2,530 individuals who clustered with the individuals in 1000G with EUR, AFR and ASN ancestries respectively. We removed a total of 652 individuals indicated as having "putative.sex.chromosome.aneuploidy" ("Indicates samples identified as putatively carrying sex chromosome configurations that are not either XX or XY")[1], as well as 71,143, 542 and 70 individuals with EUR, AFR and ASN ancestries respectively with relatedness greater than 0.05 ('Kinship' score in UKBiobank full release sample QC file) with any of the 337,127 individuals with White-British ancestries in UKbiobank we use in our main analyses, or to each other. We therefore retain 51,567, 7,497 and 2,460 unrelated individuals of EUR, AFR and ASN ancestries for our analyses. For each group, we performed PCA with all 5,327,974 biallelic SNPs using flashPCA[8], and showed that they contain few individuals of discordant self-reported ancestries. We use the top 20 PCs from each group as covariates for all following analyses.

For PRS analysis, we used 2,441,319, 1,808,453 and 2,025,285 SNPs (MAF >= 0.05, INFO score >= 0.9, P value for HWE violation > 10-6) in the EUR, ASN and AFR individuals in UKBiobank respectively, and calculated PRS for the three phenotypes (deep, EHR and EDGAR liability) for each of the nine diseases using PRSice v2[9], using the options --clump-kb 250kb --clump-p 1 --clump-r2 0.1 --interval 5e-05 --lower 5e-08. We used the top 20 genomic PCs from the EUR, ASN and AFR individuals in UKBiobank as covariates to control for population structure in each of the cohorts.

### EDGAR model

#### *Hyperparameter tuning*

We adopted the same hyperparameter settings as in the previous AutoComplete study[10]. Specifically, we employed copy masking with a masking ratio of 0.8, a learning rate of 0.1, an encoding ratio of 1, a model depth of 1, and trained for 100 epochs. The batch size was set to 4,096 to fully utilize the available memory on the NVIDIA RTX 4090 GPU.

#### *Comparison with previous models*

This study presents, to our knowledge, the first systematic framework for predicting lifetime disease liability across multiple disease domains in the UKBiobank, leveraging both EHR diagnostic codes and disease-relevant measures and trained with deep clinical phenotypes as gold-standard labels. Previous methods have not relied heavily on disease relevant measures nor used deep-clinical phenotypes to assess prediction accuracy[11,12]. Consequently, no pre-existing benchmarks are directly applicable to this specific prediction goal.

#### *Feature contributions measured by SHAP values*

To maximize predictive accuracy and clinical utility, our integrated models incorporate all available phenotypic data, including the biological measures involved in deep phenotype definitions. We emphasize that these definitions are multi-faceted; for example, labels for DBT, HLDL, and HTN incorporate self-reported medication data (excluded from model inputs), while GAD and MDD are derived from complex diagnostic criteria involving multiple symptomatic components (**Supplementary Table S2**).

To characterize the underlying predictive logic, we obtain SHAP values[13,14] from the corresponding prediction models (EHR + relevant features, red bar in **Figure 1c**). We show full results in **Supplementary Figures S1-9**. The most important features include not only the biomarkers involved in deep phenotype definition, but also the corresponding EHR code and key demographic information (age, sex). This distribution confirms that the model successfully aligns fragmented clinical records with continuous biological measures to estimate a latent disease liability, rather than performing a trivial mapping of a single diagnostic threshold.

We perform targeted feature ablations to evaluate how the model behaves when primary diagnostic markers are unavailable. By removing the defining features for ANEM (Haemoglobin concentration, hb), COPD (FEV1/FVC ratio Z-score and FEV1 predicted percentage) and NAFLD (Alanine aminotransferase, alt), we effectively test the model's ability to recover the disease signal from the remaining data. We find that this results in a limited decrease in prediction accuracy (ANEM: AUC 0.98→0.76; COPD: AUC 0.99→0.95; NAFLD: AUC 1.00→0.97), demonstrating the model’s ability to leverage biological redundancy; upon removal of a primary feature, the model automatically pivots to highly correlated physiological proxies (e.g., AST in the absence of ALT for NAFLD). These results, together with simulation for realistic feature availability, demonstrate the robustness in gain of using disease-relevant information across a wide range of possible scenarios, where the lower boundary lies exactly at the scenario when there are only EHR features.

### Transformer model

#### *Comparison with Delphi*

We have provided the Delphi[15] performance as a reference to compare with our EDGAR model, as both utilize UKBiobank EHR data. However, our models diverge in two important ways: prediction objective and model architecture.

For the prediction objective, our model estimates lifetime disease liability—defined as the propensity for one to develop disease over one's lifetime. In our model we assume the most deeply phenotyped definition of the disease, obtained from the UKBiobank (**Supplementary Table S2**), captures this disease liability, and we therefore use it as a label for model training. As the deeply phenotyped definition of disease is obtained using different approaches from EHR diagnostic codes, we do not expect it to capture exactly the same biases EHR diagnostic codes do, and therefore using them as labels enables our predictions to move away from biases specific to the EHR. In contrast, Delphi's prediction objective is a future disease episode, predicting the probability of clinical events within a future time window based on existing EHR codes. As it only learns from EHR events (episodes of the same or different diseases), we expect it to also learn the inherent biases in the EHR, such that the predictions of future disease episodes encompass those biases.

Architecturally, our EDGAR model is a tabular multi-layer perceptron that works on tabular counts of EHR diagnostic codes for different diseases, whereas Delphi utilizes a transformer architecture designed to process longitudinal sequences of discrete EHR events. To evaluate whether our tabular architecture is as good as a transformer architecture to capture relevant information from EHR events for liability prediction, we implemented an encoder-only transformer baseline that replicates the core sequential architecture and self-supervised paradigm of Delphi.

In this baseline model, each EHR event is represented by the sum of three embeddings: a code embedding (reflecting sequence of ICD-10 codes), a time embedding (reflecting age at event), and a positional embedding (reflecting sequence order). The resulting representations are passed through a multi-layer transformer encoder with multi-head self-attention to model temporal and contextual relationships among diagnoses. The model is trained under joint supervision for both next-token prediction and disease liability prediction. Next-token prediction is similar to the way Delphi was trained, using a masked language modeling (MLM) head that reconstructs masked diagnosis codes to encourage temporal and contextual learning. The disease liability prediction is a classification head predicting multiple deep phenotype labels when available. The total loss combines MLM and classification losses with a tunable weight $\alpha$, allowing the model to learn both general EHR representations (like Delphi) and disease liabilities. We perform hyperparameter tuning on the validation split to optimize the model prediction.

#### *Sequence of ICD-10 codes*

Following prior work[15,16] that treats diagnostic codes as word tokens[17] in longitudinal EHR sequences, we first construct a unified vocabulary of diagnosis codes across HES (ICD-9/10) and GP (Read v2/v3) records by mapping all codes to the ICD-10 standard (3-digit level, e.g., S618) where possible, resulting in a final vocabulary of 10,730 unique tokens, 17,248,752 total events (tokens) for 316,968 patients (White British). Each patient's events are chronologically ordered by age at diagnosis, and every unique code is assigned a token ID along with special tokens [PAD], [MASK], [UNK], [Sex: Female], [Sex: Male].

#### *Hyperparameter tuning*

The search space included embedding dimension d_model $\in \{64, 128, 256\}$, number of Transformer layers n_layers $\in \{3, 4, 6\}$, attention heads n_heads $\in \{2, 4\}$, loss weighting coefficient $\alpha \in \{0.5, 1.0\}$, and masking ratio mask_ratio $\in \{0.2, 0.4\}$. All experiments use a batch size of 128, learning rate = $1 \times 10^{-3}$, maximum sequence length = 128, and 60 training epochs for consistent comparison. Performance is monitored on a validation set (20% of training data) after each epoch using macro-AUC, with early stopping applied after 10 epochs of non-improvement. The final model configuration is selected based on the highest validation macro-AUC, and the corresponding checkpoint is subsequently evaluated on the held-out test set. All experiments are conducted on a single NVIDIA RTX 4090 GPU.

### Active learning

#### *Active learning methods*

In a budget-limited patient call-back scenario, active learning[18] is a well-studied paradigm that directly addresses this label acquisition bottleneck by optimizing the querying priority — that is, deciding which unlabeled individuals should be prioritized for call-back. Two major operational regimes are commonly used: stream-based (online) querying[19], where samples arrive sequentially and the model decides on-the-fly whether to query their labels; and pool-based (batch) querying, where the model iteratively selects a subset of samples from a fixed unlabeled pool for labeling in batches. The latter is more suitable for patient call-back prioritization, as the former is prohibitively expensive and operationally impractical both on the computational and the hospital logistics ends.

Within the pool-based (batch) active learning setting, two main families of approaches are commonly used: a) diversity-based methods like Coreset[20], which select representative samples to maximize feature-space coverage; and b) uncertainty-based methods like Conf[21], which prioritize samples predicted with the least confidence. Hybrid methods attempt to balance diversity and uncertainty to maintain robustness across batch sizes, often through a tunable weighting parameter. The state-of-the-art approach, Badge[18] is a notable exception: it is a parameter-free hybrid method that embeds samples in the gradient space, thereby jointly capturing uncertainty and representativeness.

#### *Applications in supervised vs semi-supervised learning*

Although active learning has been extensively studied[18,20–22], it is traditionally applied within supervised learning settings. In such settings, models are trained only on labeled data selected by the active learning module, while unlabeled data remain unused until annotated. In contrast, semi-supervised learning models like AutoComplete[3] can still leverage unlabeled data through self-supervised objectives such as masked language modeling (MLM), improving generalization even before any new labels are acquired. Consequently, diversity-based active learning methods[20] may be less effective in semi-supervised settings, as the unlabeled data already provide coverage of the input distribution that diversity sampling aims to achieve. As a result, selecting diverse samples provides limited additional benefit, and may even introduce samples that are less informative for refining the decision boundary.

We see this in our study, where Coreset[20] performs worse than random sampling in some instances (**Figure 2b**). This observation supports the hypothesis that uncertainty-driven selection better complements semi-supervised learning, where the primary gain from new labels lies in correcting uncertain or ambiguous regions rather than improving representativeness of the training set.

**External GWAS**

We assembled two sets of external GWAS for validation: (i) External Deep phenotype–based GWAS and (ii) External EHR-based GWAS (**Figure 4a**). External EHR GWAS summary statistics were obtained from FinnGen (data freeze DF12) using matched ICD-10 codes (**Supplementary Table S7**). For diabetes, we used type II diabetes (T2D) as the external EHR phenotype, as a combined T1D/T2D GWAS is not available in FinnGen, and our DBT Deep phenotype shows no genetic correlation with FinnGen T1D (rG = 0.04, SE = 0.07, p = 0.51).

For External Deep GWAS (**Supplementary Table S8**), strictly matched phenotype definitions are generally unavailable in existing studies. We therefore obtained the most biologically proximal GWAS that capture core disease-relevant mechanisms. Specifically, for seven non-psychiatric disorders, we used biomarker-based GWAS, including hemoglobin[23] (ANEM), lung function[24] (FEV1/FVC; COPD), hemoglobin A1c[23] (DBT), LDL cholesterol[25] (HLDL), diastolic blood pressure[23] (HTN), alanine transaminase[23] (NAFLD), and femoral neck bone mineral density[26] (OST).

For psychiatric disorders, no external GWAS precisely matches our GAD definition; most available studies[27,28] include broader anxiety-related conditions such as panic and phobias. We therefore used summary statistics from an anxiety factor derived via genomic structural equation modeling across multiple cohorts[29]. For MDD, recent GWAS[30] incorporates heterogeneous measurement sources, including clinical diagnoses, EHR, questionnaires, and self-reports. We selected the GWAS restricted to clinically ascertained cases, which aligns more closely with deep phenotype definitions, despite its smaller effective sample size (effective sample size Neff = 62,534) compared with the mixed-source GWAS (Neff = 967,078)[30]. The heterogeneity in phenotype definitions[31,32], and limited statistical power in clinical MDD phenotype is likely to explain why GWAS hits for MDD from EDGAR liabilities show higher replication in External EHR GWAS than in External Deep GWAS (**Figure 4c**). External Deep GWAS for GAD includes partial sample overlap with UK Biobank, reflecting data availability, whereas the remaining external GWAS are non-overlapping.

All external GWAS summary statistics were lifted over to the GRCh37 reference genome. For each genome-wide significant locus identified in the discovery GWAS ($p < 5 \times 10^{-8}$), we defined replication as the presence of at least one SNP within ±250 kb of the lead variant that reached significance in the external GWAS at $p < 0.05$, Bonferroni-corrected for the total number of discovery loci.

### Bias identification and removal

#### *Methodological considerations for bias identification*

We define 'disease-specific bias' as the genetic component of the EHR phenotype that is independent of the EDGAR liability for each specific disease. We further define the 'Common Bias' as the shared factor underlying all disease-specific biases. We apply a GWAS-by-subtraction[33] framework to isolate each disease-specific bias (**Figure 5d**) and then use a common factor model to extract the Common Bias (**Figure 5g**). Our bias identification framework treats the EDGAR liability as an approximation of true disease liability. We incorporate two primary methodological considerations regarding this approach.

First, if the EDGAR liability contains systematic biases or miscalibrations—arising from model architecture, training data, or subtle ascertainment effects—these components will be partitioned into the latent liability factor rather than the bias factor. This would lead to a more conservative estimate of bias, potentially underestimating its true magnitude. However, this limitation implies that the biases we report likely represent lower bounds, ensuring the robustness of our findings. We note that the Common Bias is unlikely to reflect any bias present in the deep phenotype. Taking potential participation bias in the deep phenotypes as an illustrative example: If our EDGAR liability inherits participation bias in the deep phenotype, then participation bias will be partitioned into the disease liability factor (**Figure 5b**), and EHR-specific bias will be independent of it (due to orthogonality restraints, **Figure 5b).** As a result, the Common Bias factor will be irrelevant to the participation bias in the deep phenotypes. Its genetic correlation with participation bias (**Figure 5i**) reflects EHR-specific bias.

Second, a potential challenge in disease-specific bias identification arises if the EDGAR liability fails to fully capture the etiological signal present in the corresponding EHR phenotype (e.g., as observed with OST). In such cases, uncaptured etiological effects may "spill over" into the latent bias factor. To mitigate this risk, we leverage a Common Factor model to derive a systemic bias component (**Figure 5g**). By aggregating bias signals across diverse diseases, our framework selectively extracts shared confounders while isolating and discarding disease-specific residual etiological effects. This hierarchical approach ensures that the resulting Common Bias factor remains a robust representation of systemic EHR noise.

#### *Alternative model to Common Bias: Psychiatric Bias and Non-Psychiatric Bias*

We test an alternative model with two bias factors: a psychiatric bias factor (biasing MDD and GAD) and a non-psychiatric bias factor (biasing the seven non-psychiatric diseases). Although the model fit well ($\chi^2$=31.37, df=26, p=0.21, AIC=69.37, CFI=0.99, SRMR=0.09, **Supplementary Figure S28**), it is not significantly better than the single Common Bias model ($\chi^2$ difference test, p = 0.18). Thus, we can not reject the hypothesis of a single common confounder. We also test the rG between these two bias factors with socioeconomic/behavior traits (**Supplementary Table S10)**. The non-psychiatric bias largely mirrored the original Common Bias in its correlation patterns (**Supplementary Figure S29, Supplementary Table S10)**. However, the psychiatric bias showed distinct correlations; specifically, it differed significantly from non-psychiatric bias regarding BMI, number of illnesses, loneliness, being ever a smoker, neuroticism, and deprivation index (**Supplementary Figure S29,**). While this suggests the psychiatric bias may be distinct, the fact that it does not provide a significantly better factor model fit prevents us from conclusively favoring the two-factor model over the Common Bias model.

#### *Bias removal*

The goal of the bias removal model is to remove the common confounder identified in **Figure 5g** from an external EHR GWAS. A formal specification of the model is shown in **Figure 6b**. To achieve this, we apply the GWAS-by-Subtraction model to external EHR GWAS and Common Bias, by subtracting the latent bias factor ($g_{Bias}$) from EHR ($EHR_{ex}$). To assess whether this improves rG with true disease liability, we introduce a disease liability factor ($Liab_{in}$), observed from our EDGAR liability phenotype.

#### *Mathematical Proof of Increased Genetic Correlation via Bias Removal*

We aim to prove that removing the latent bias component from a GWAS on an external EHR phenotype ($EHR_{ex}$) theoretically guarantees an increase in its genetic correlation (rG) with our EDGAR disease liability in UKBiobank ($Liab_{in}$), under the assumption that the latent systematic bias is orthogonal to the true disease-specific liability.

Let the genetic component of the external EHR phenotype, $g_{EHR_{ex}}$, be modeled as a linear combination of the true latent liability ($g_{Liab_{ex}}$) and the latent bias ($g_{Bias}$), as illustrated in **Figure 6b**:

$$g_{EHR_{ex}} = g_{Liab_{ex}} + \gamma \cdot g_{Bias},$$

where $\gamma \geq 0$, and the bias is independent of the true disease liability:

$$Cov(g_{Liab_{ex}}, g_{Bias}) = 0$$

The EDGAR liability of the same disease in UKBiobank ($g_{Liab_{in}}$) acts as an unbiased anchor. It is correlated with the true external liability but independent of the latent systematic bias.

$$Cov(g_{Liab_{in}}, g_{Bias}) = 0$$
$$Cov(g_{Liab_{in}}, g_{Liab_{ex}}) = \sigma_{ie} > 0$$

Before bias removal, the $rG_{before}$ (between $EHR_{ex}$ and $Liab_{in}$) can be written as

$$rG_{before} = r_g(EHR_{ex}, Liab_{in}) = \frac{Cov(g_{EHR_{ex}}, g_{Liab_{in}})}{\sqrt{Var(g_{EHR_{ex}}) \cdot Var(g_{Liab_{in}})}}$$

Substituting $g_{EHR_{ex}} = g_{Liab_{ex}} + \gamma \cdot g_{Bias}$ into the numerator:

$$\begin{aligned} Cov(g_{EHR_{ex}}, g_{Liab_{in}}) &= Cov(g_{Liab_{ex}} + \gamma g_{Bias}, g_{Liab_{in}}) \\ &= Cov(g_{Liab_{ex}}, g_{Liab_{in}}) + \gamma \cdot Cov(g_{Bias}, g_{Liab_{in}}) \\ &= Cov(g_{Liab_{ex}}, g_{Liab_{in}}) \\ &= \sigma_{ie} \end{aligned}$$

Thus,

$$rG_{before} = \frac{\sigma_{ie}}{\sqrt{Var(g_{EHR_{ex}}) \cdot Var(g_{Liab_{in}})}}$$

After bias removal, the $rG_{after}$ (between $Liab_{ex}$ and $Liab_{in}$) can be written as

$$rG_{after} = r_g(Liab_{ex}, Liab_{in}) = \frac{\sigma_{ie}}{\sqrt{Var(g_{Liab_{ex}}) \cdot Var(g_{Liab_{in}})}}$$

Since the numerator remains constant ($\sigma_{ie}$), the difference between $rG_{before}$ and $rG_{after}$ is driven exclusively by the reduction in phenotypic genetic variance after removing the additive bias component:

$$Var(g_{EHR_{ex}}) = Var(g_{Liab_{ex}} + \gamma g_{Bias}) = Var(g_{Liab_{ex}}) + \gamma^2 Var(g_{Bias}) \geq Var(g_{Liab_{ex}})$$

We can prove that:

$$rG_{after} \geq rG_{before}$$

**Factor associations with socioeconomic and behavioral traits**

We estimate genetic correlation between each one of the four latent factors with 43 socioeconomic and behavioral traits[4,34–38]. The four latent factors include the Common Bias factor, the Common EHR factor, and the Psychiatric-Bias and Non-Psychiatric-Bias factors from the alternative model. The 43 traits are selected due to their potential relevance to health-related behavior and are listed in **Suppmentary Table S13**. For interpretability, the traits can be grouped into four broad categories: 1) Educational attainment (EA)-related traits, including age when finished full-time education, reaction time, and educational choices. 2) General health, mental distress, and lifestyle traits. These include BMI, number of illnesses, cancer, insomnia, broad depression and anxiety, dietary intake, physical activity and smoking behaviors. 3) Participation behaviors, risking behaviors and the Big Five personalities. 4) Sociodemographics, including deprivation index and sex. We apply Bonferroni correction to the p-values of rG for a total number of 172 tests.

**Supplementary Figures**

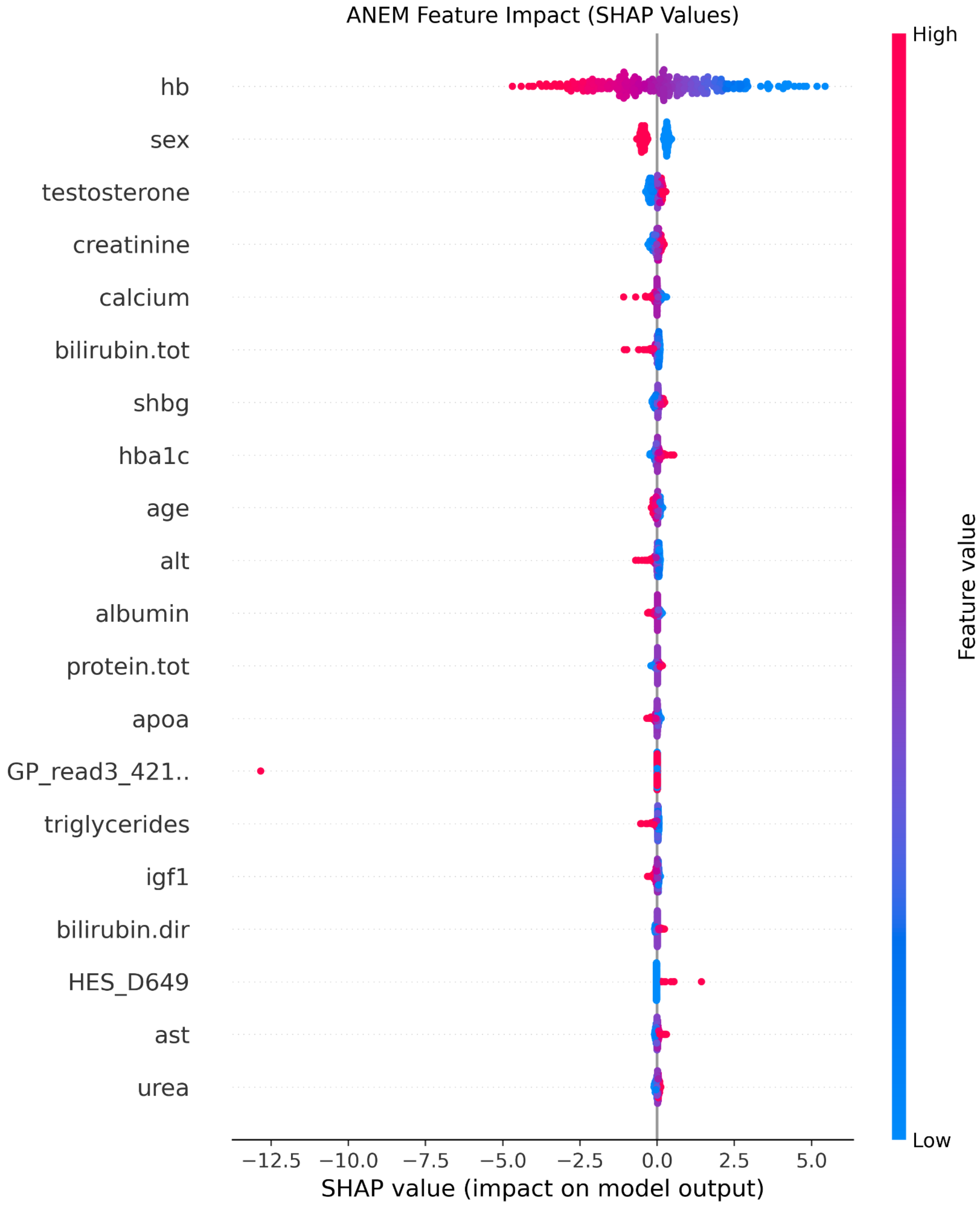

**Supplementary Figure S1.** SHAP value for predicting ANEM. GP_read3_421..: Haematology - general. HES_D649: Anaemia, unspecified.

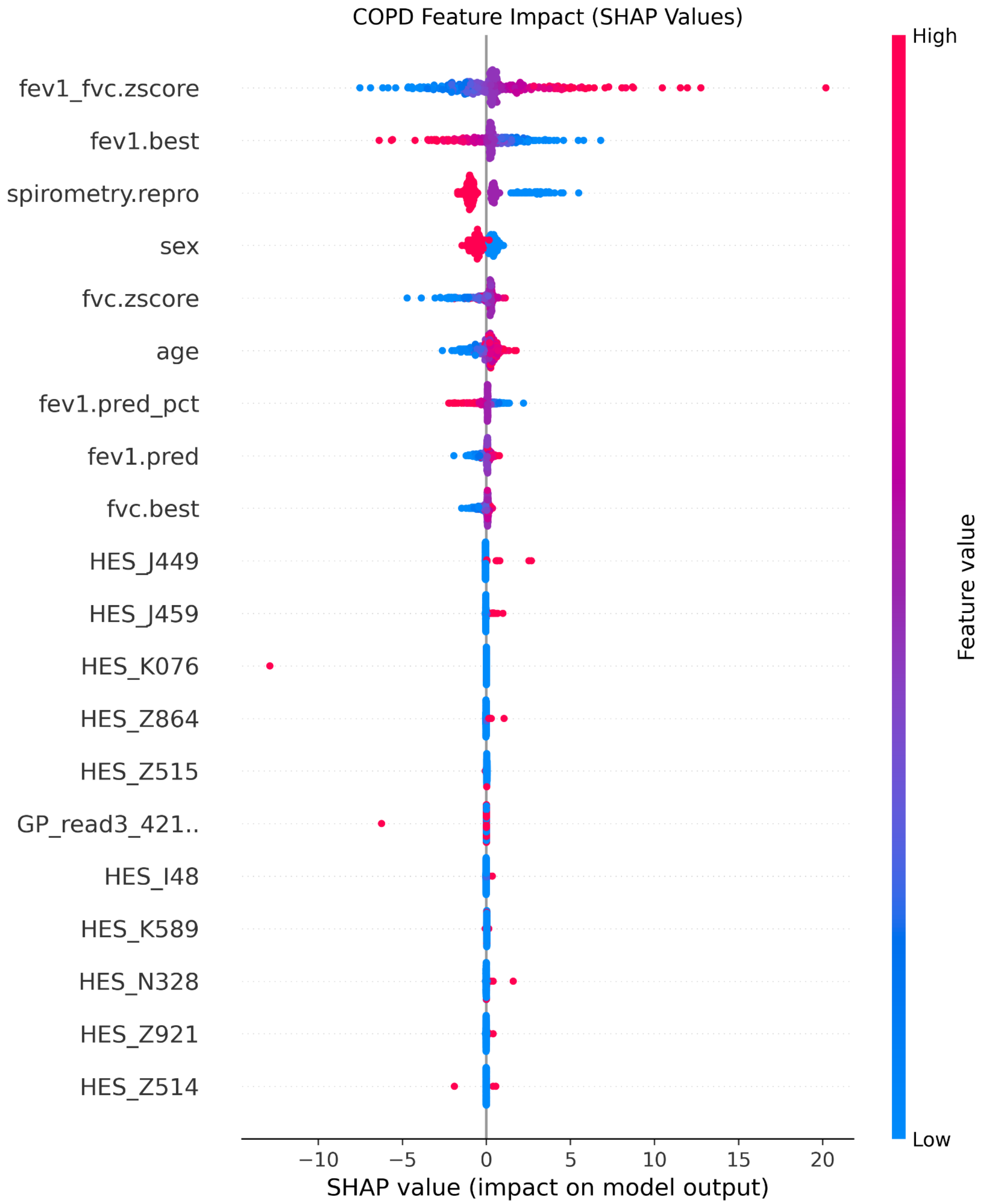

**Supplementary Figure S2.** SHAP value for predicting COPD. HES_J449: Chronic obstructive pulmonary disease, unspecified. HES_J459: Asthma, unspecified. HES_K076: Temporomandibular joint disorders. HES_Z864: Personal history of psychoactive substance abuse. HES_Z515: Palliative care. GP_read3_421..: Haematology - general. HES_I48: Atrial fibrillation and flutter. HES_K589: Irritable bowel syndrome without diarrhoea. HES_N328: Other specified disorders of bladder. HES_Z921: Personal history of long-term (current) use of anticoagulants. HES_Z514: Preparatory care for subsequent treatment, not elsewhere classified.

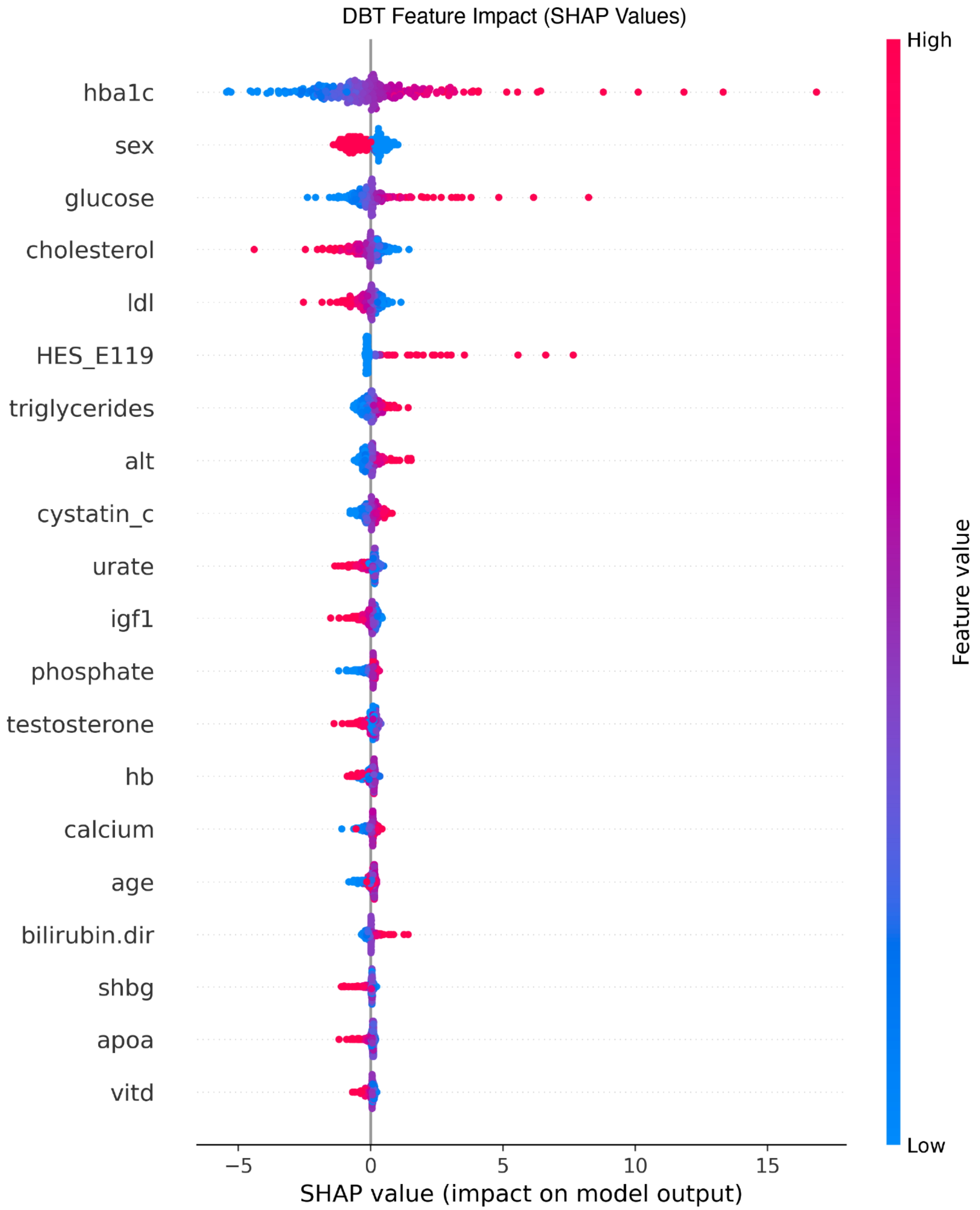

**Supplementary Figure S3.** SHAP value for predicting DBT. HES_E119: Type 2 diabetes mellitus.

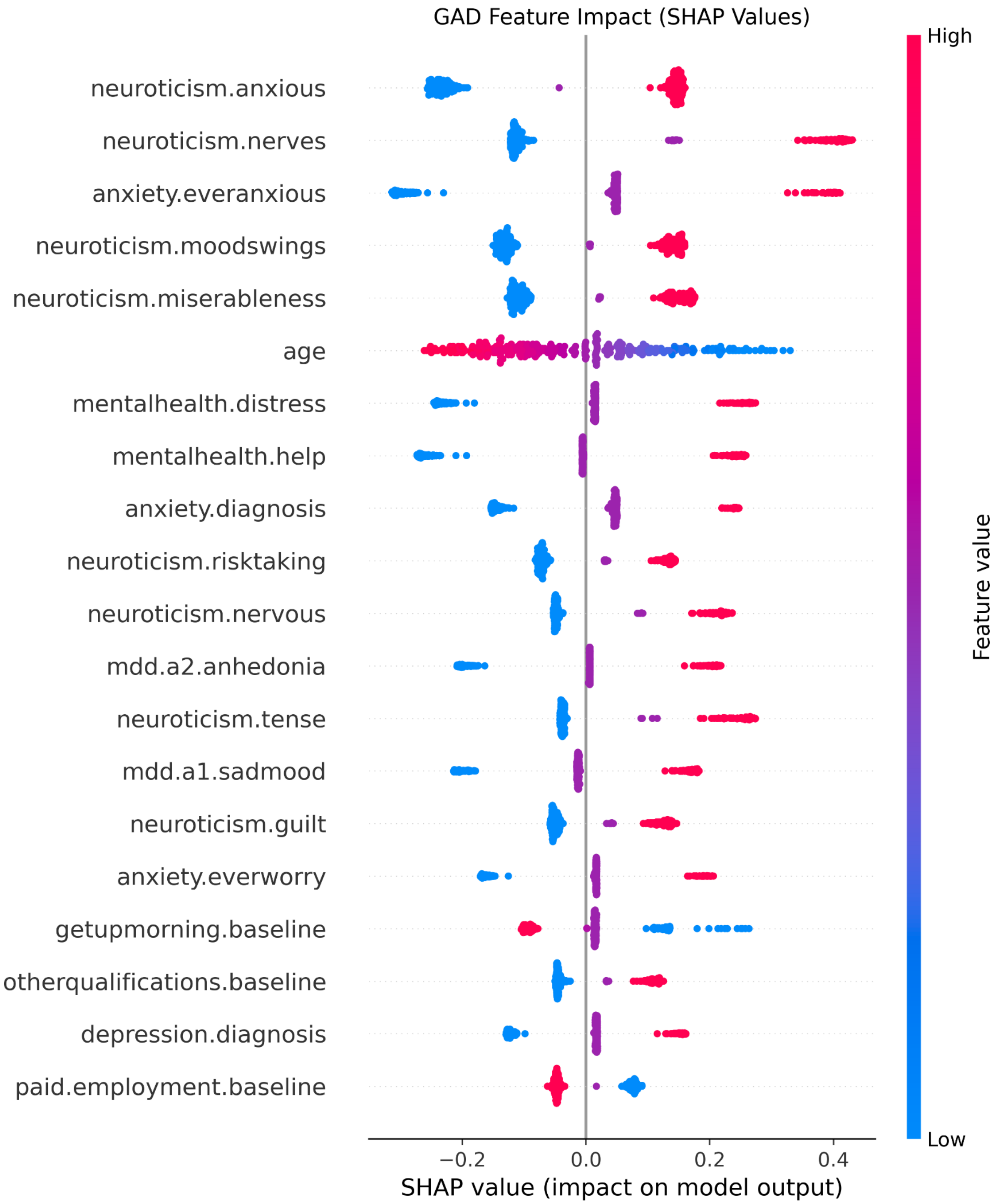

**Supplementary Figure S4.** SHAP value for predicting GAD.

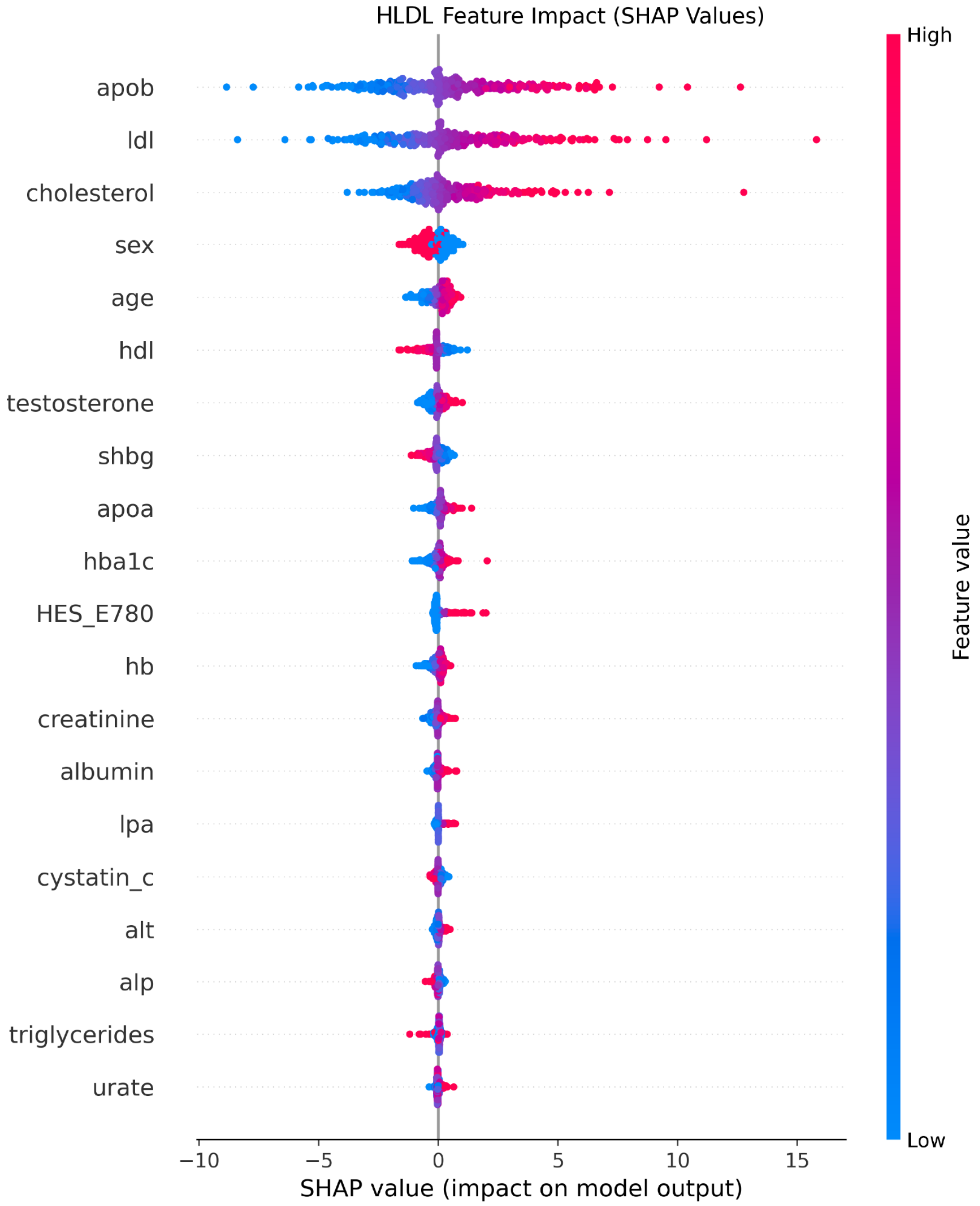

**Supplementary Figure S5.** SHAP value for predicting HLDL. HES_E780: Pure hypercholesterolaemia.

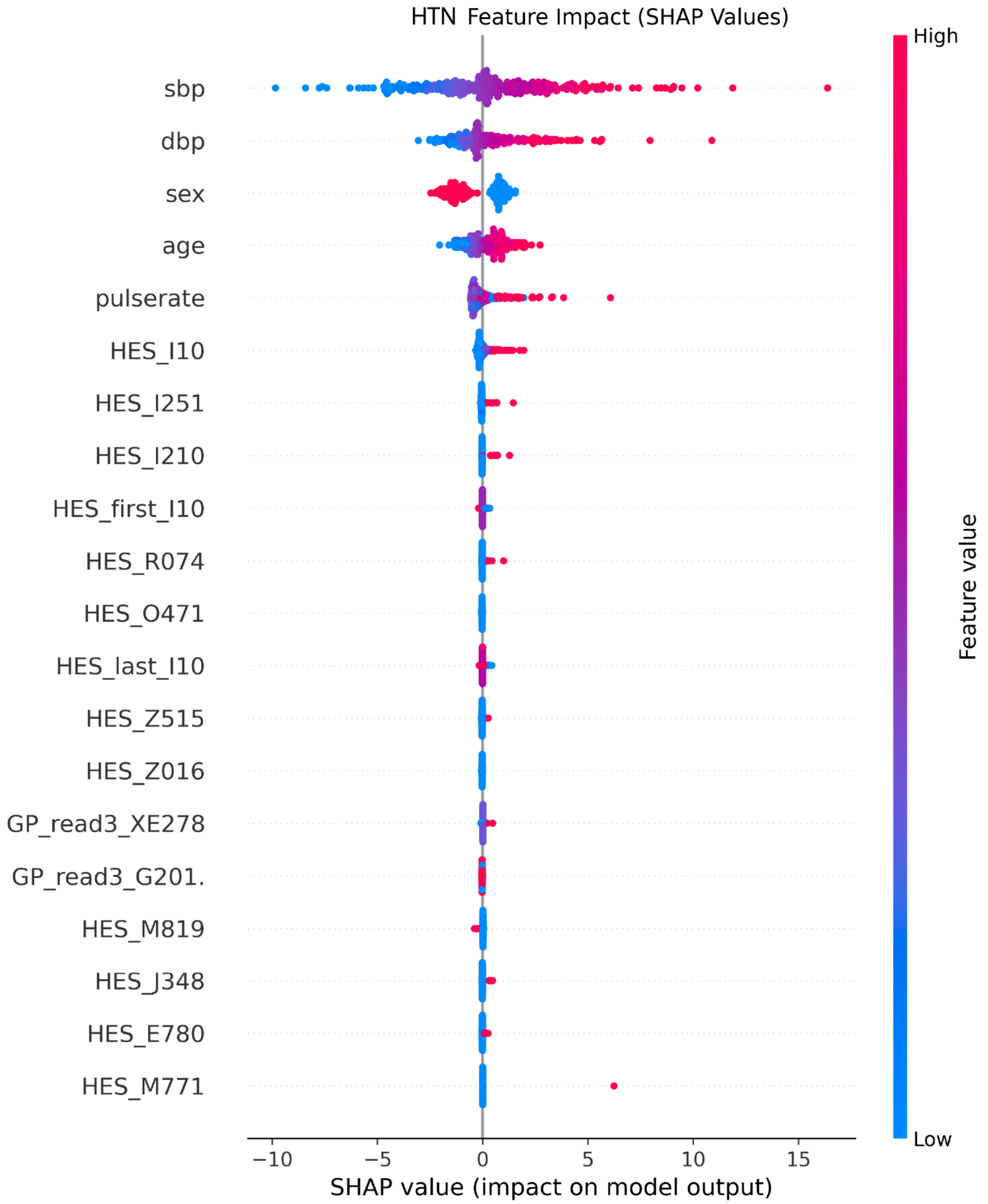

**Supplementary Figure S6.** SHAP value for predicting HTN. HES_I10: Essential (primary) hypertension. HES_I251: Atherosclerotic heart disease. HES_I210: Acute transmural myocardial infarction of anterior wall. HES_R074: Chest pain, unspecified. HES_O471: False labour at or after 37 completed weeks of gestation. HES_Z515: Palliative care. HES_Z016: Radiological examination, not elsewhere classified. GP_read3_XE278: Cervical smear - negative. GP_read3_G201.: Benign essential hypertension. HES_M819: Osteoporosis, unspecified. HES_J348: Other specified disorders of nose and nasal sinuses. HES_E780: Pure hypercholesterolaemia. HES_M771: Lateral epicondylitis.

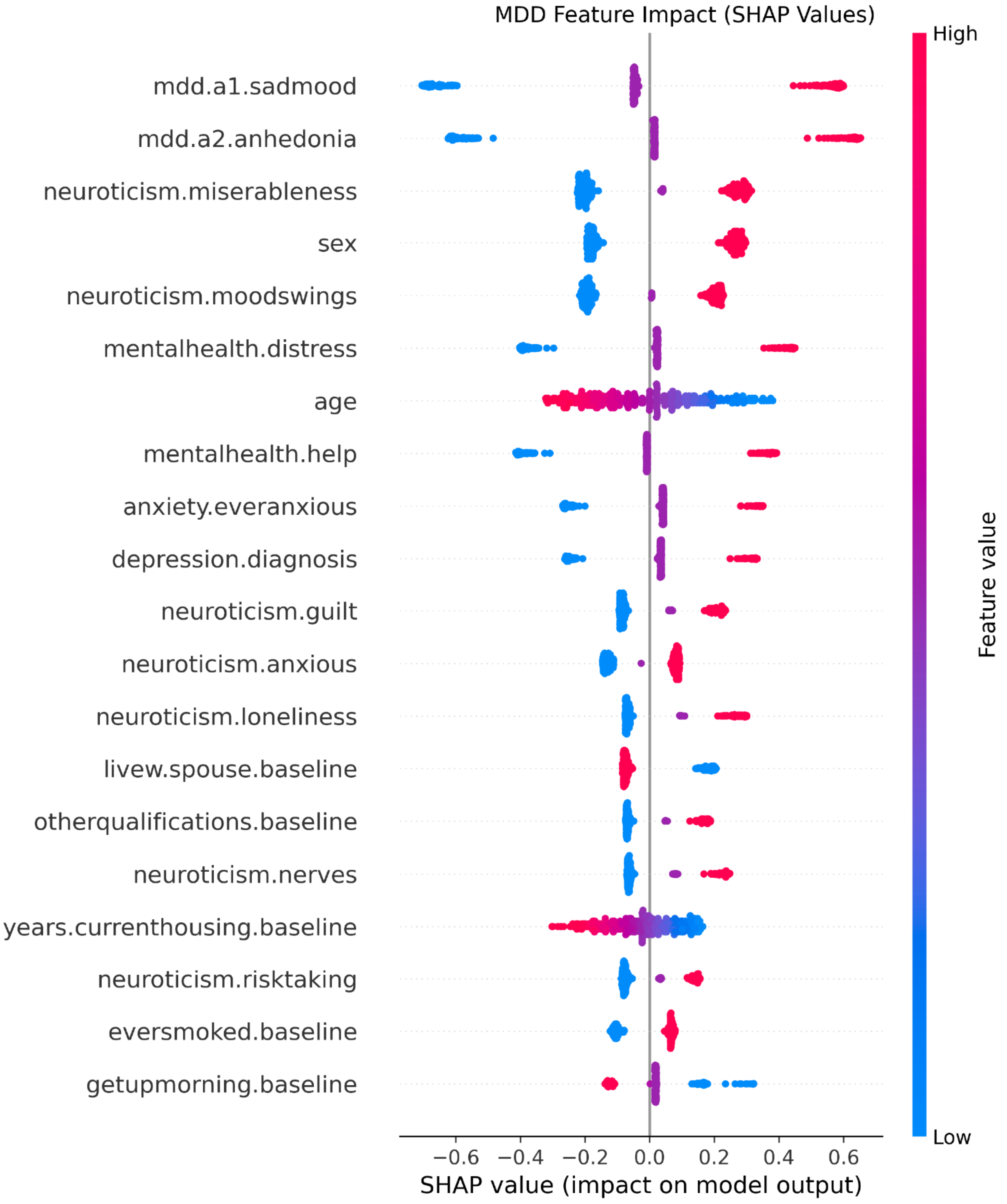

**Supplementary Figure S7.** SHAP value for predicting MDD. HES_F329: Depressive episode, unspecified.

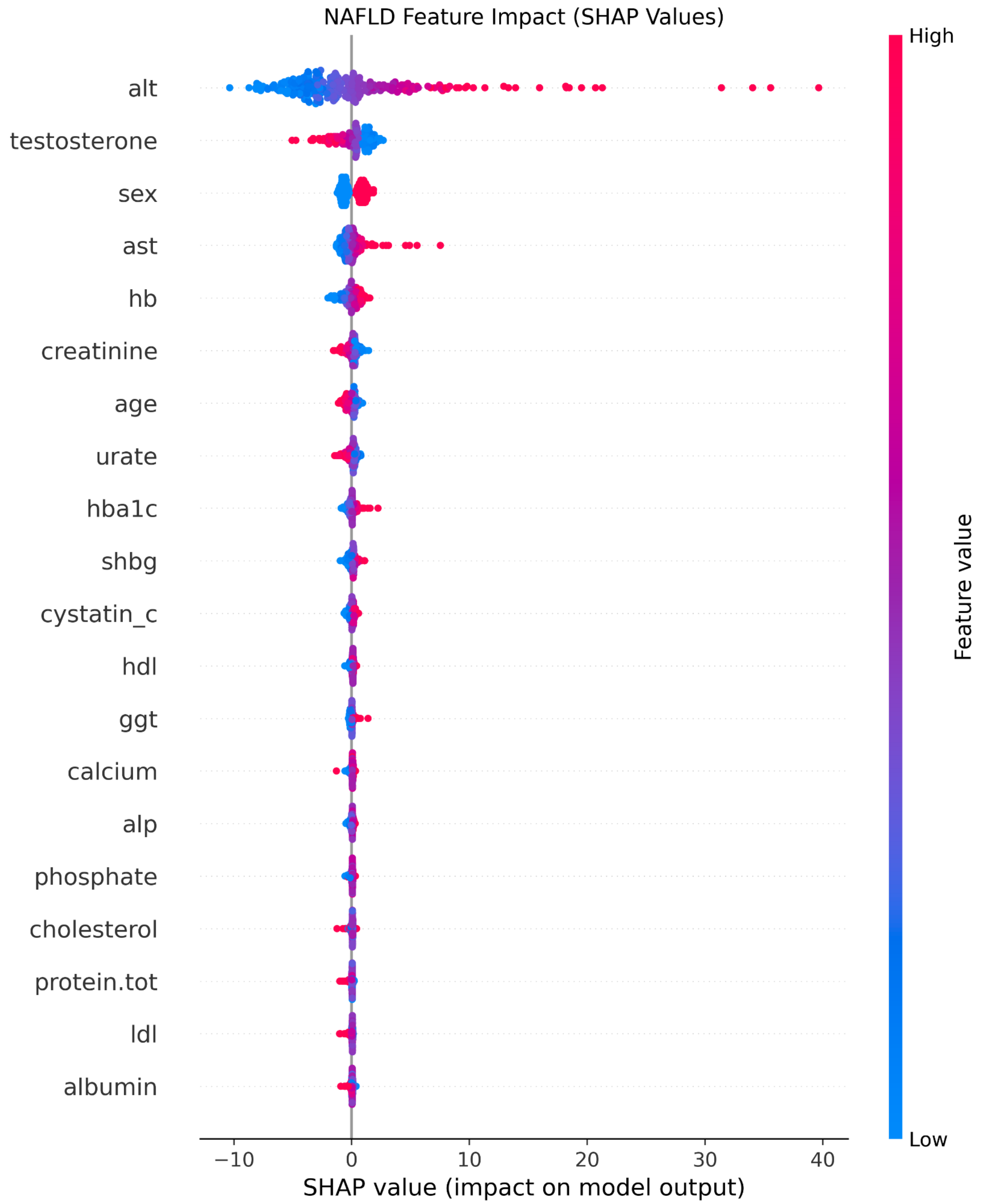

**Supplementary Figure S8.** SHAP value for predicting NAFLD.

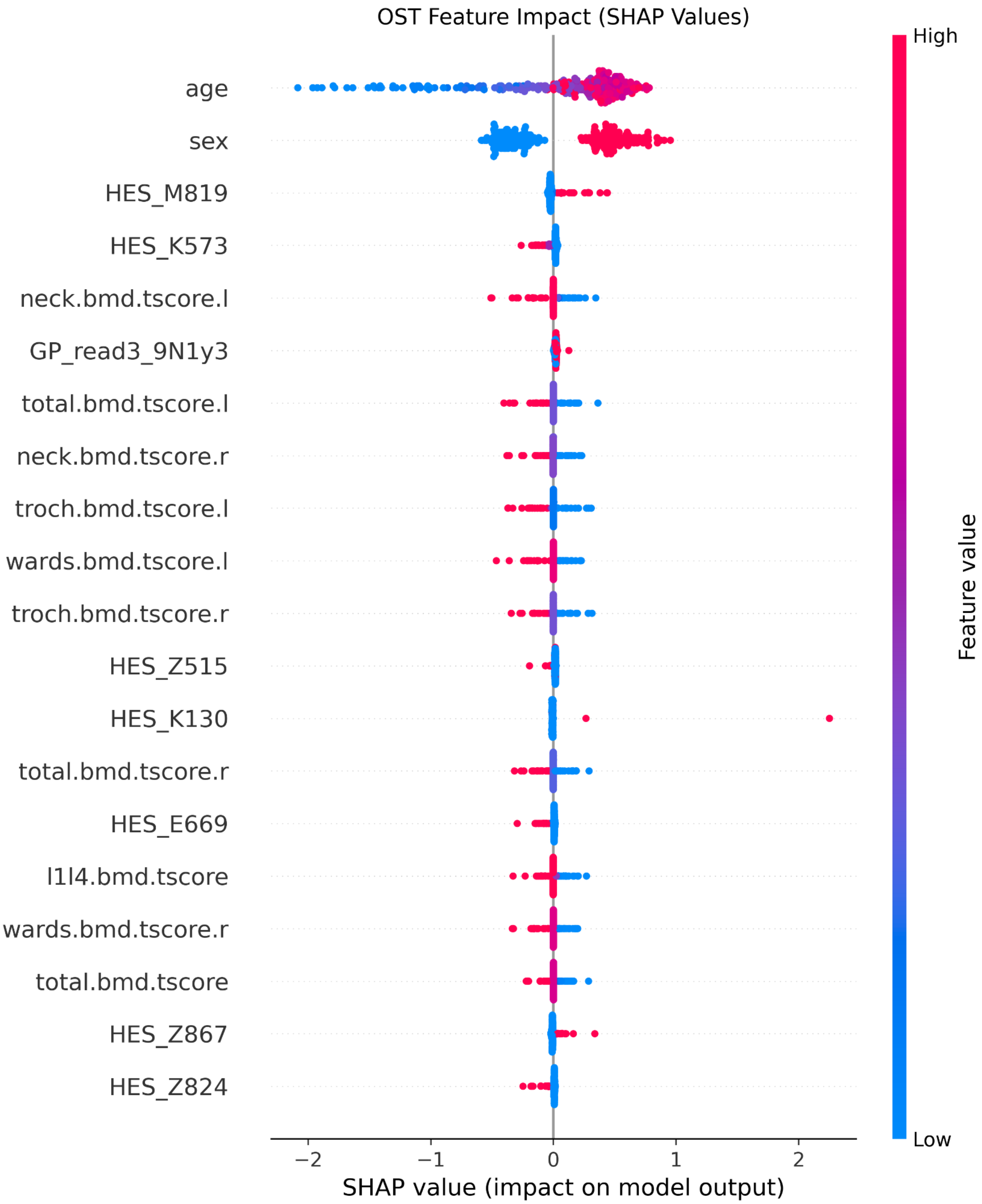

**Supplementary Figure S9.** SHAP value for predicting OST. HES_M819: Osteoporosis, unspecified. HES_K573: Diverticular disease of large intestine without perforation or abscess. GP_read3_9N1y3: Seen in emergency clinic. HES_Z515: Palliative care. HES_K130: Diseases of lips. HES_E669: Obesity, unspecified. HES_Z867: Personal history of diseases of the circulatory system. HES_Z824: Family history of ischaemic heart disease and other diseases of the circulatory system.

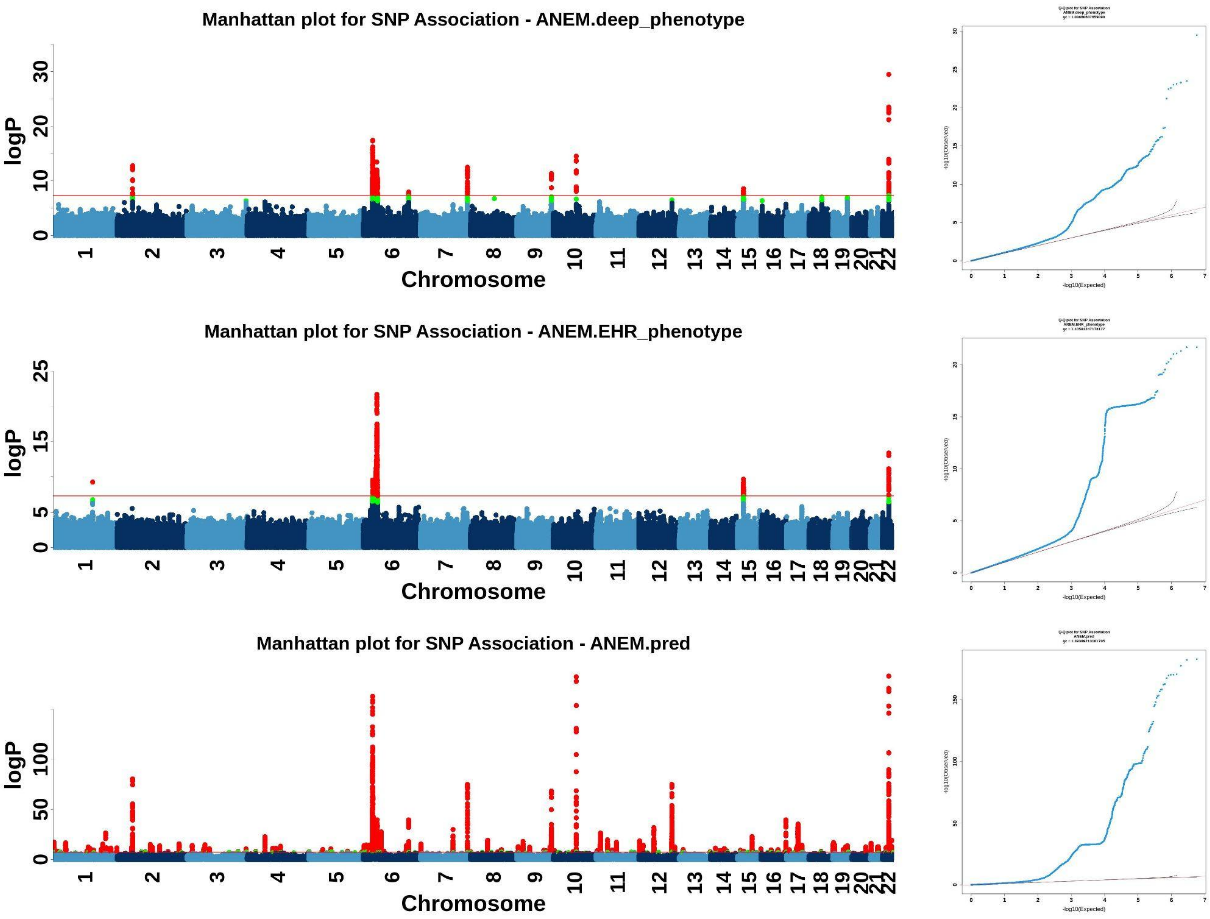

**Supplementary Figure S10.** Manhattan plots and QQ plots for ANEM across deep phenotype, EHR phenotype and EDGAR liability.

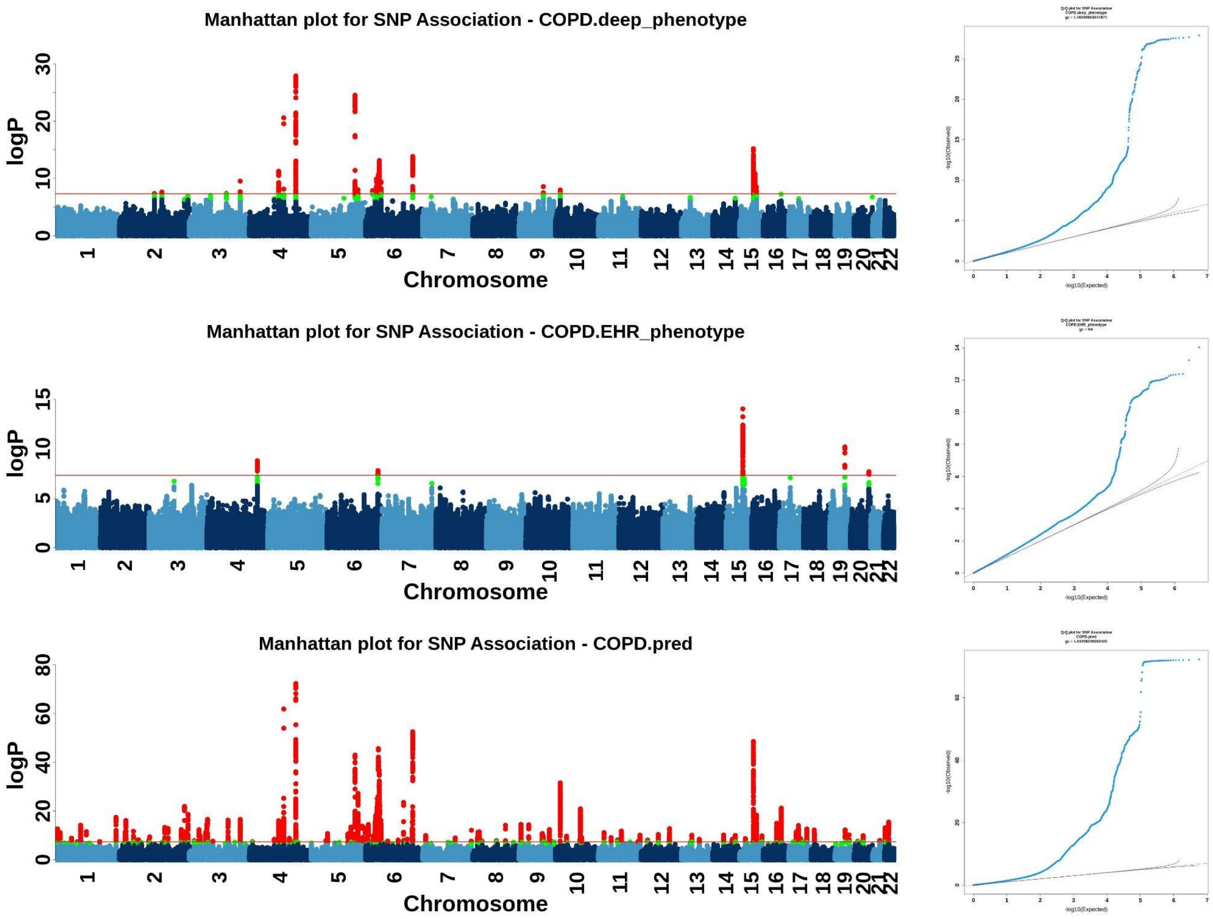

**Supplementary Figure S11.** Manhattan plots and QQ plots for COPD across deep phenotype, EHR phenotype and EDGAR liability.

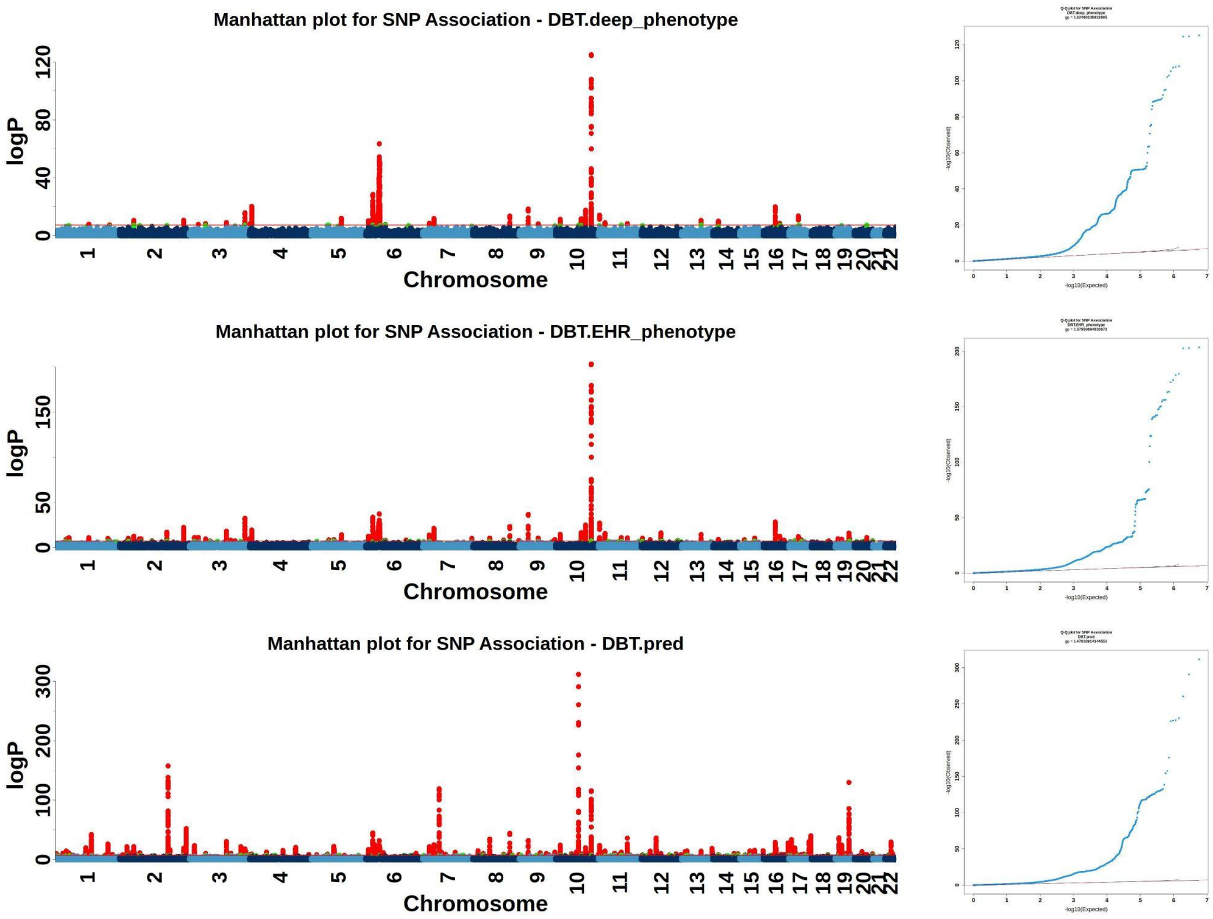

**Supplementary Figure S12.** Manhattan plots and QQ plots for DBT across deep phenotype, EHR phenotype and EDGAR liability.

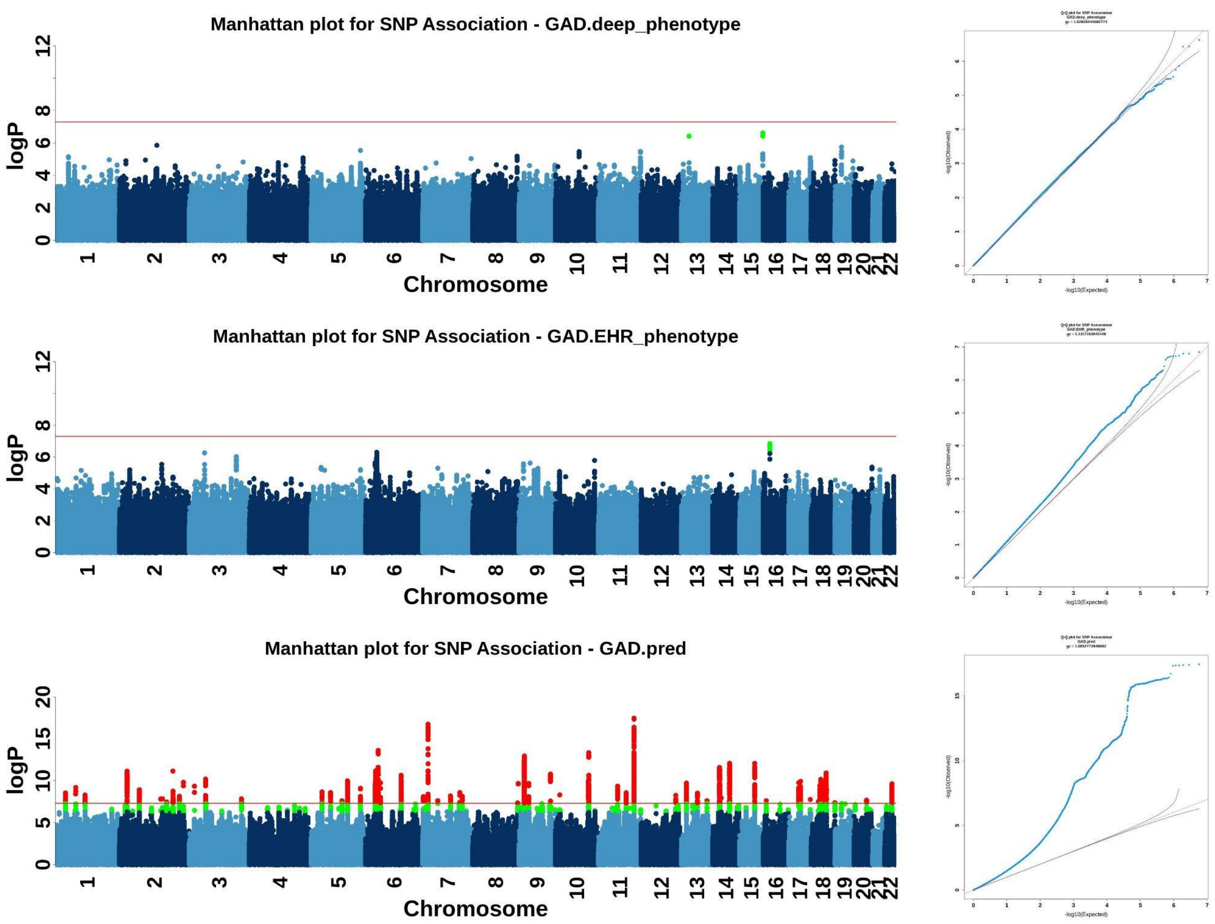

**Supplementary Figure S13.** Manhattan plots and QQ plots for GAD across deep phenotype, EHR phenotype and EDGAR liability.

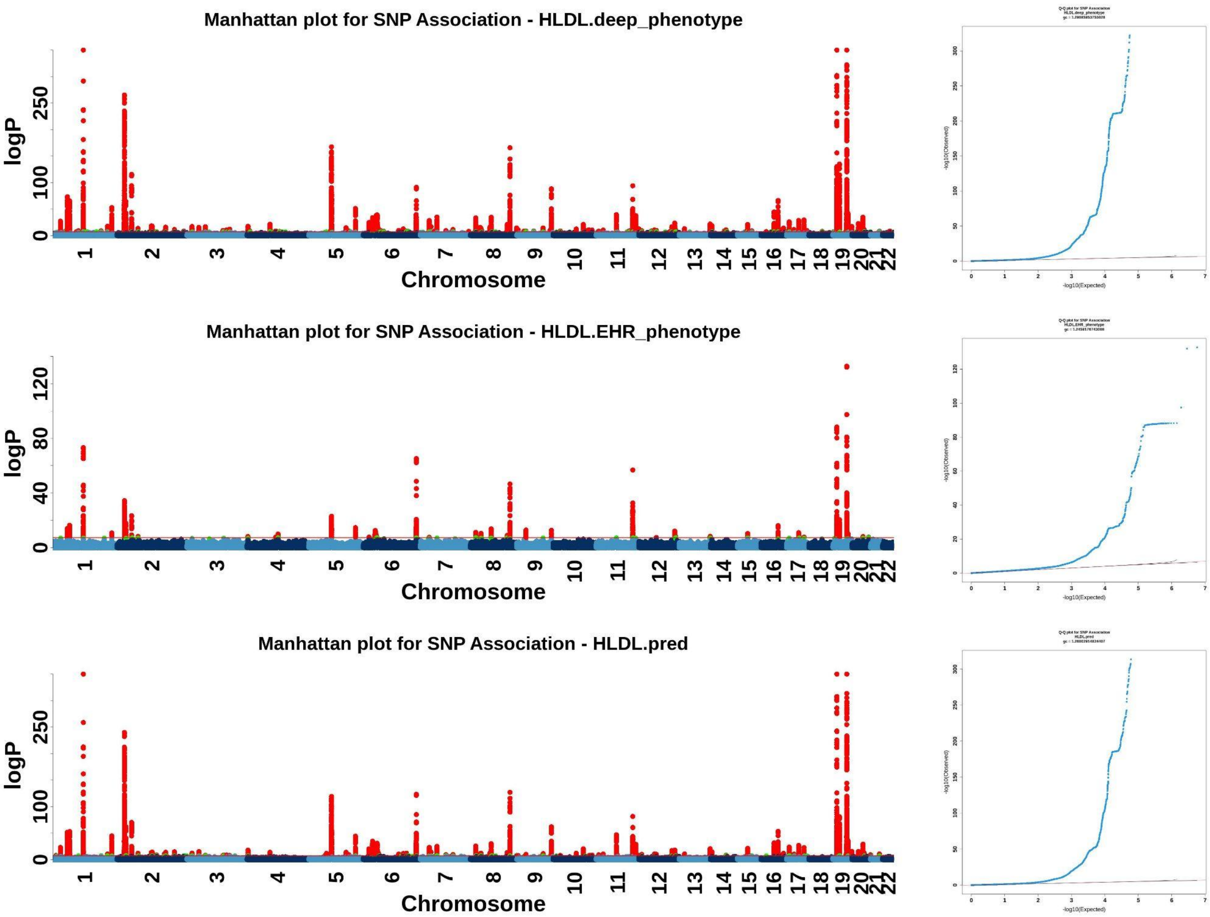

**Supplementary Figure S14.** Manhattan plots and QQ plots for HLDL across deep phenotype, EHR phenotype and EDGAR liability.

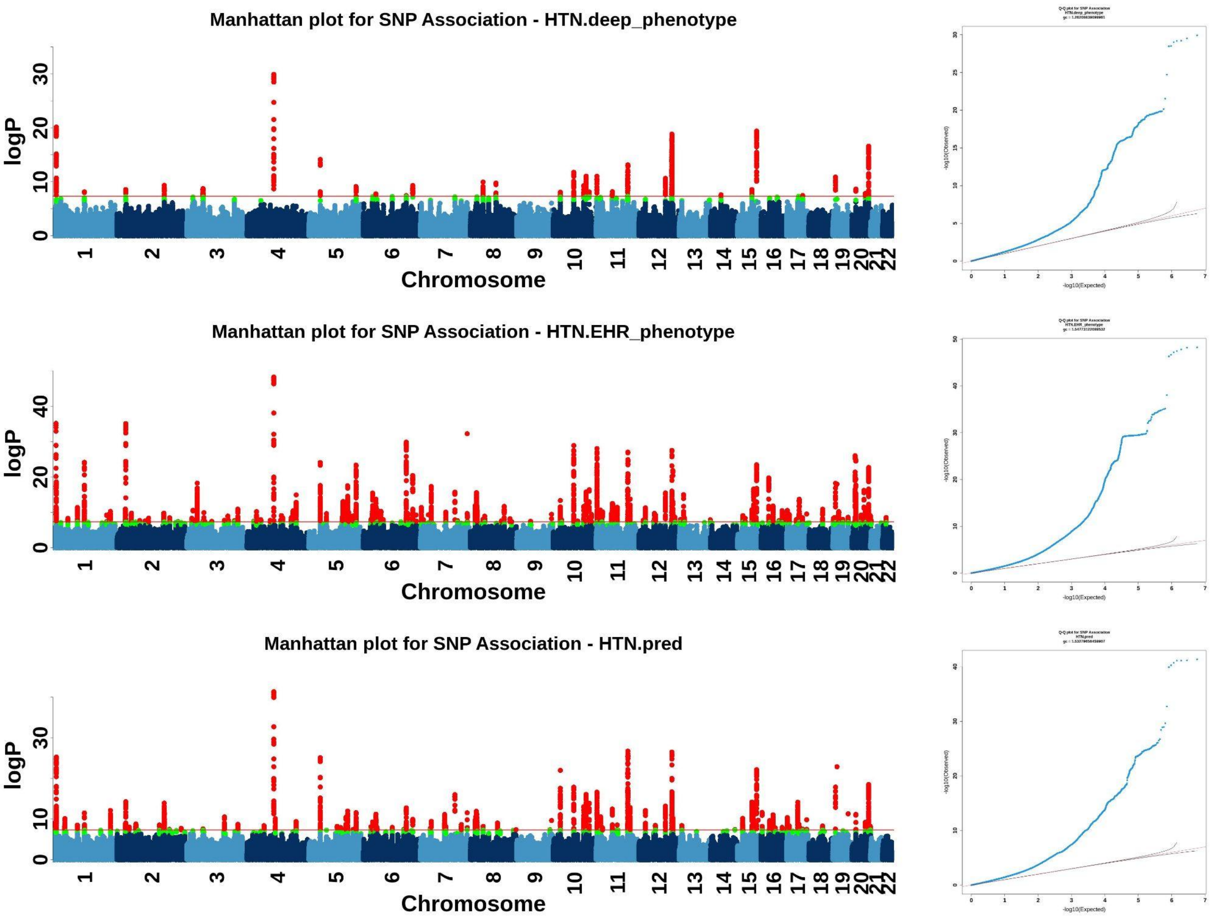

**Supplementary Figure S15.** Manhattan plots and QQ plots for HTN across deep phenotype, EHR phenotype and EDGAR liability.

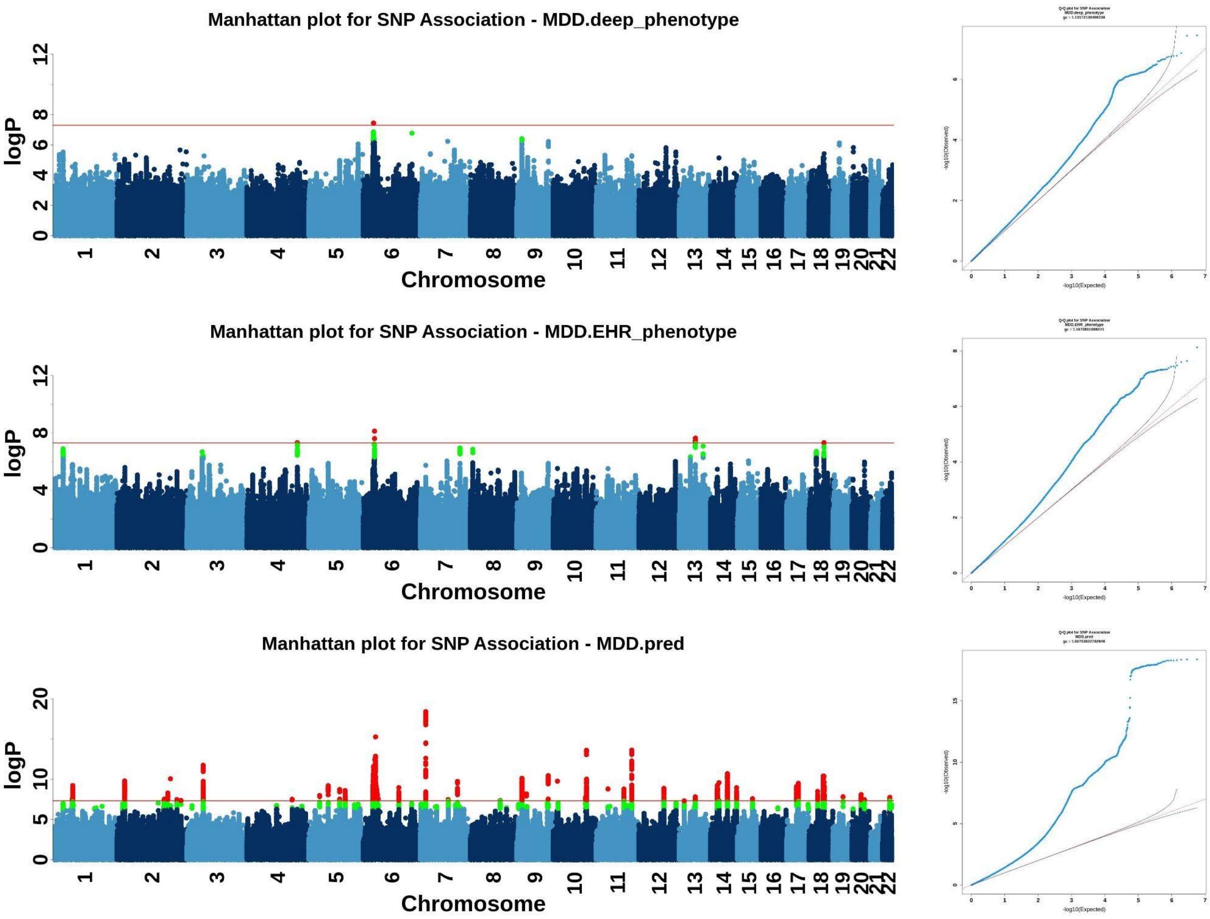

**Supplementary Figure S16.** Manhattan plots and QQ plots for MDD across deep phenotype, EHR phenotype and EDGAR liability.

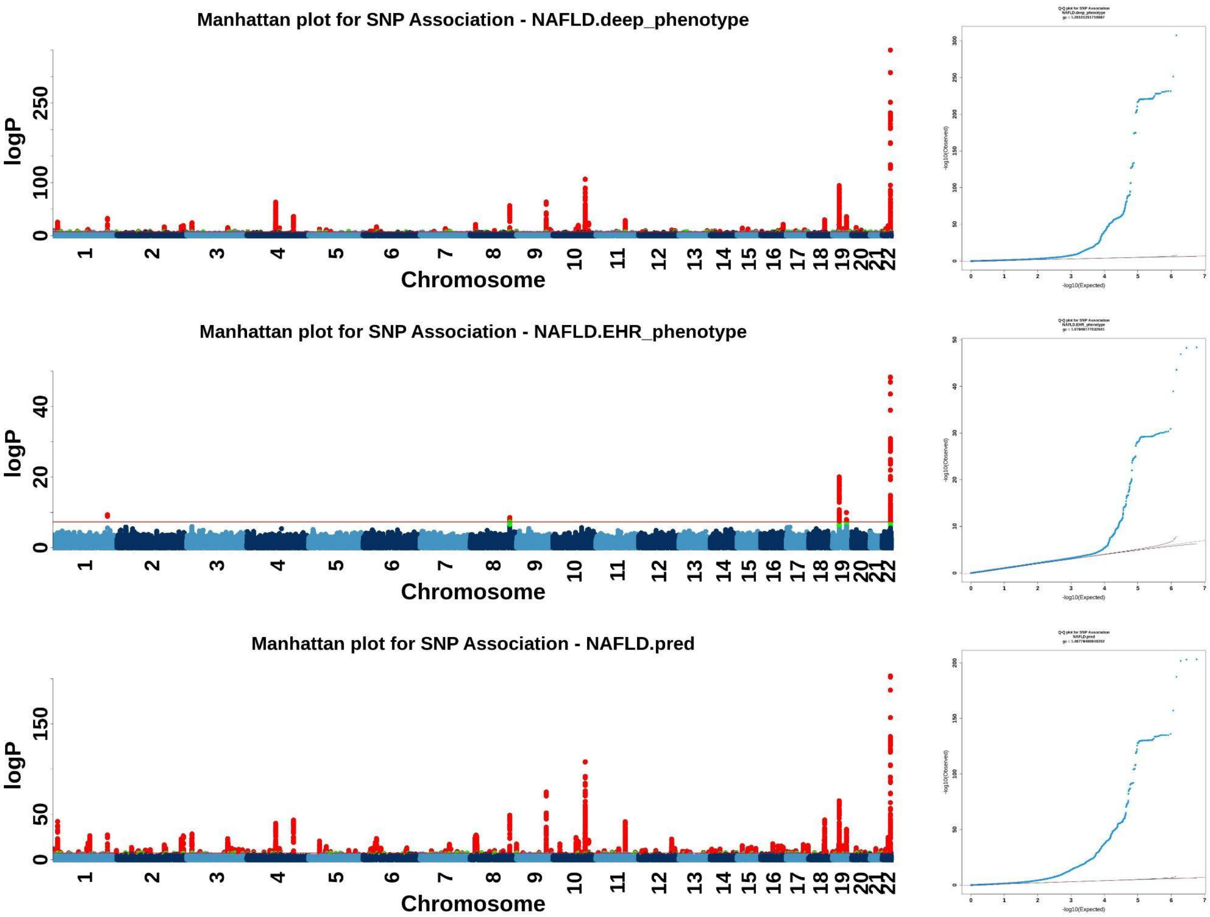

**Supplementary Figure S17.** Manhattan plots and QQ plots for NAFLD across deep phenotype, EHR phenotype and EDGAR liability.

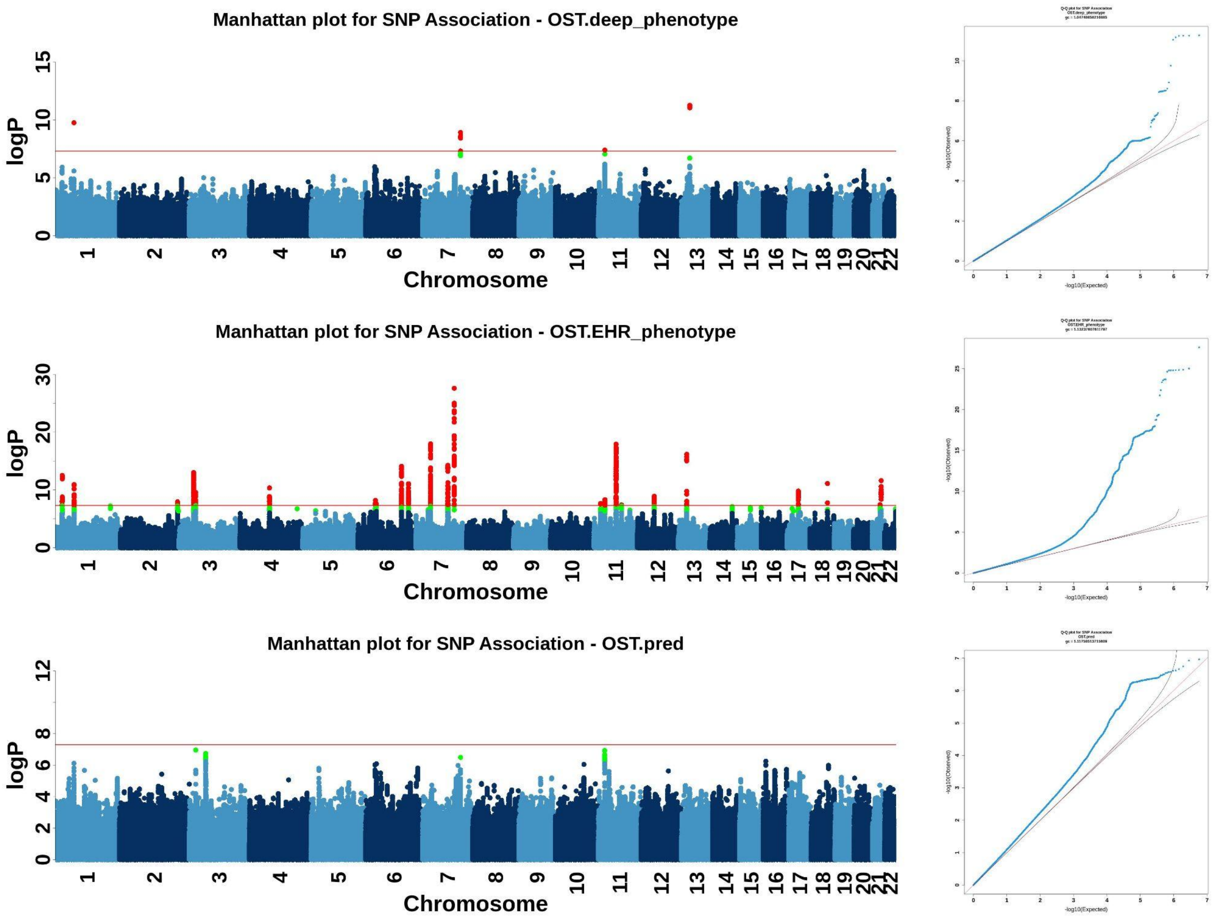

**Supplementary Figure S18.** Manhattan plots and QQ plots for OST across deep phenotype, EHR phenotype and EDGAR liability.

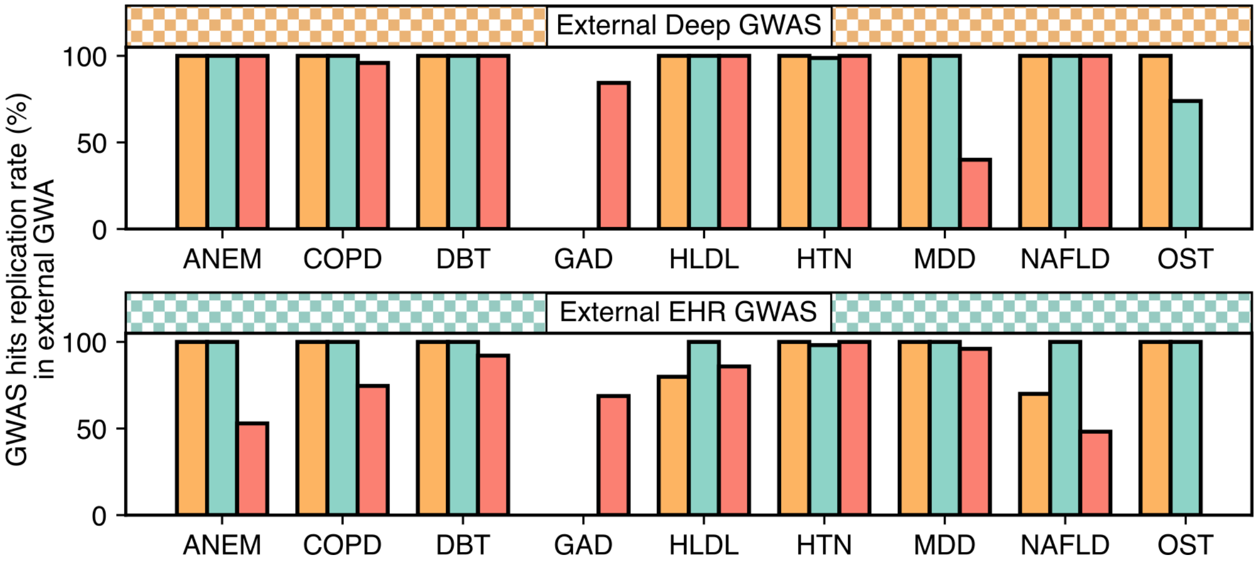

**Supplementary Figure S19. Percentage (%) of GWAS hits replicated in external GWAS.**

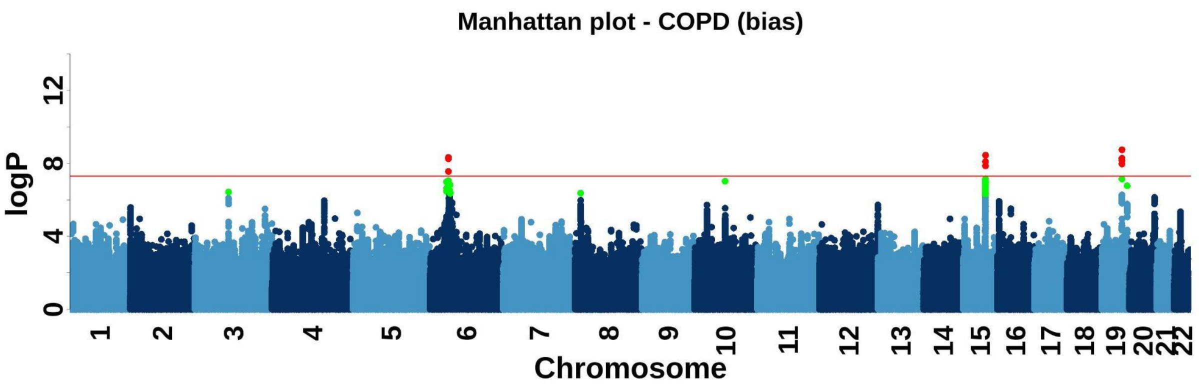

**Supplementary Figure S20.** Manhattan plots for Bias_COPD.

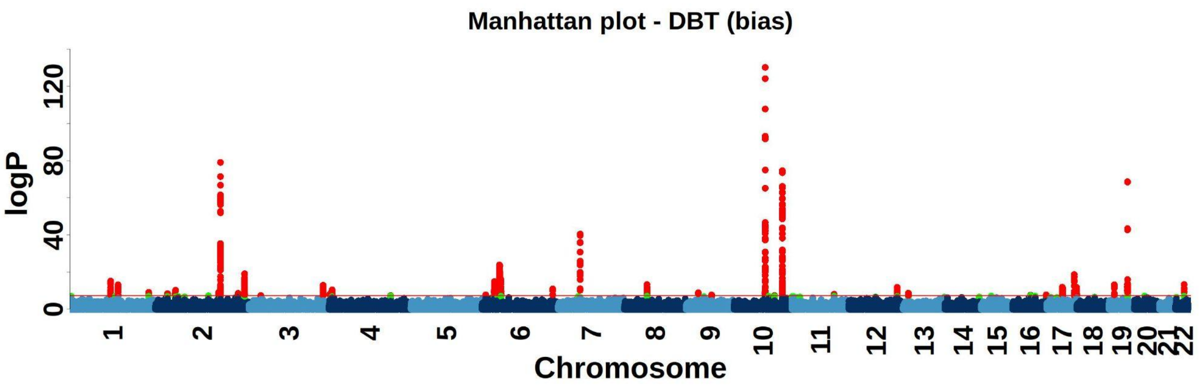

**Supplementary Figure S21.** Manhattan plots for Bias_DBT.

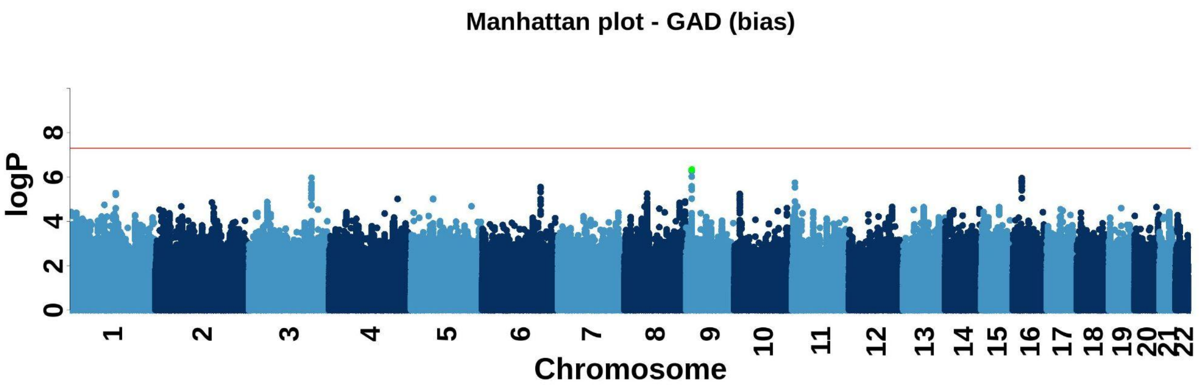

**Supplementary Figure S22.** Manhattan plots for Bias_GAD.

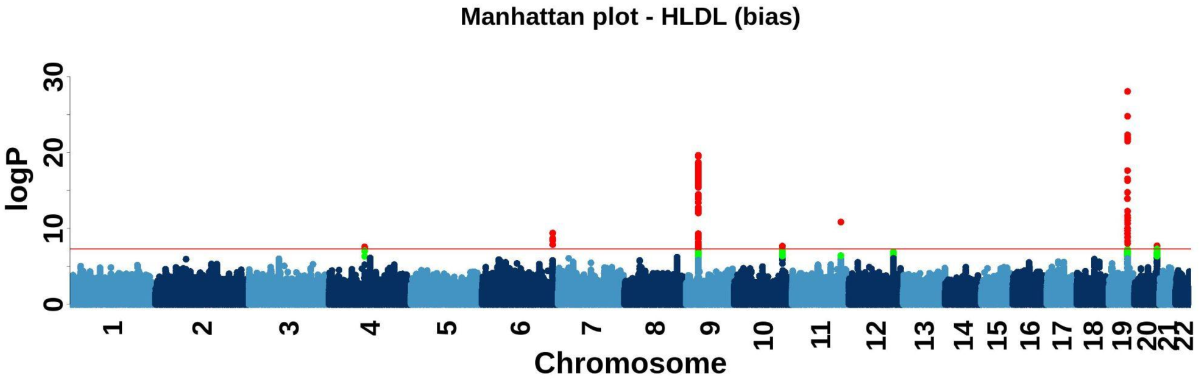

**Supplementary Figure S23.** Manhattan plots for Bias_HLDL.

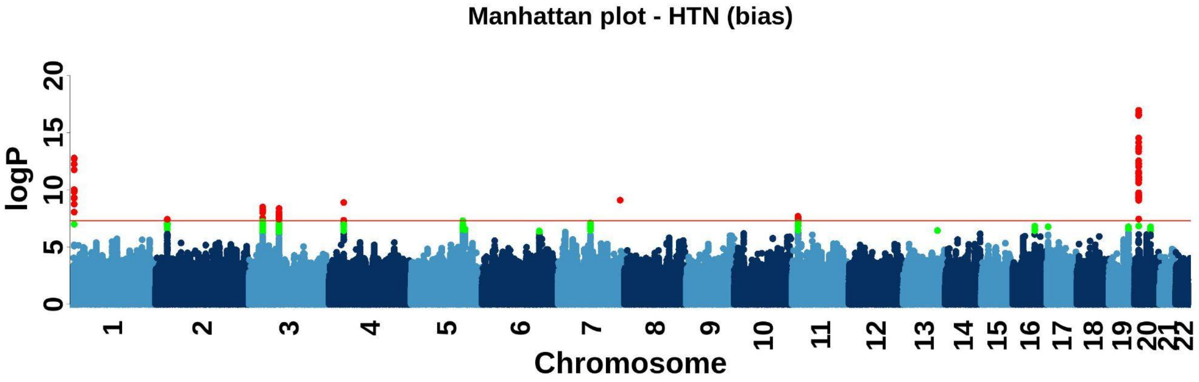

**Supplementary Figure S24.** Manhattan plots for Bias_HTN.

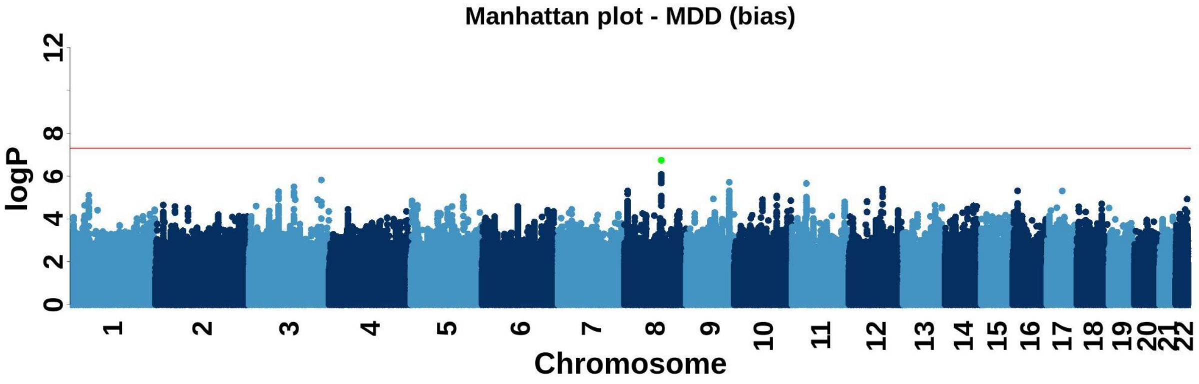

**Supplementary Figure S25.** Manhattan plots for Bias_MDD.

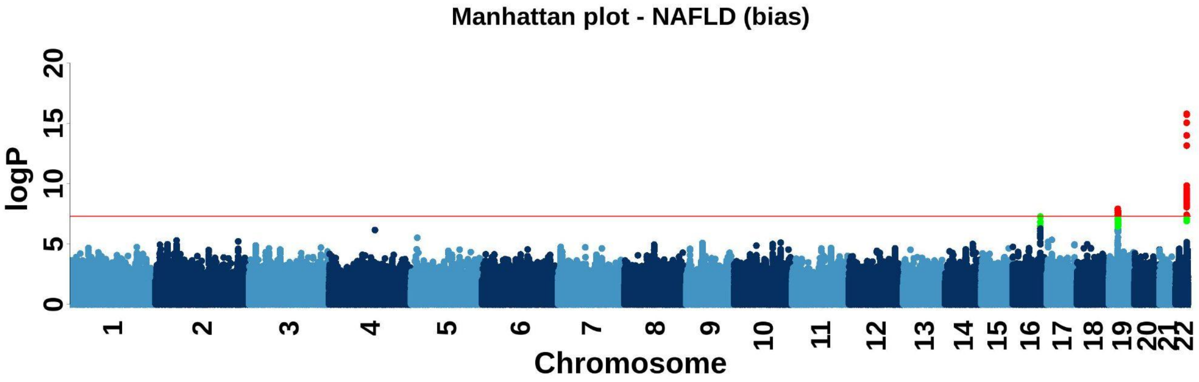

**Supplementary Figure S26.** Manhattan plots for Bias_NAFLD.

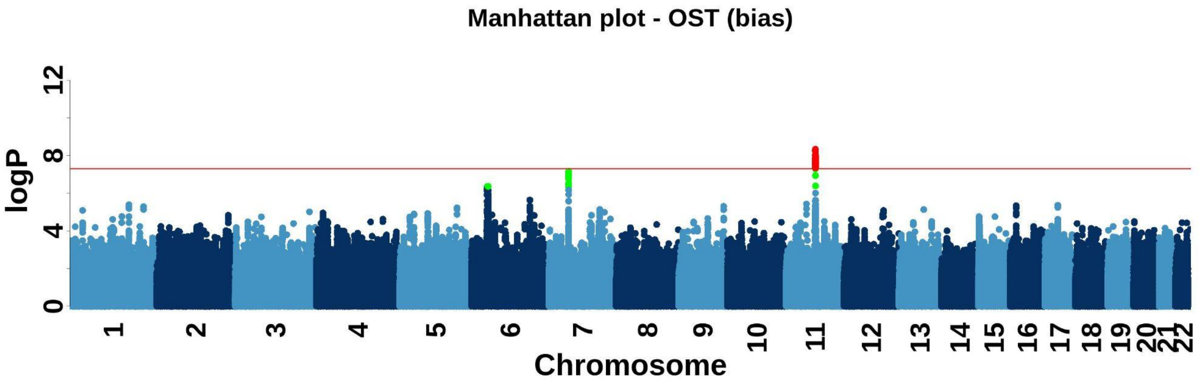

**Supplementary Figure S27.** Manhattan plots for Bias_OST.

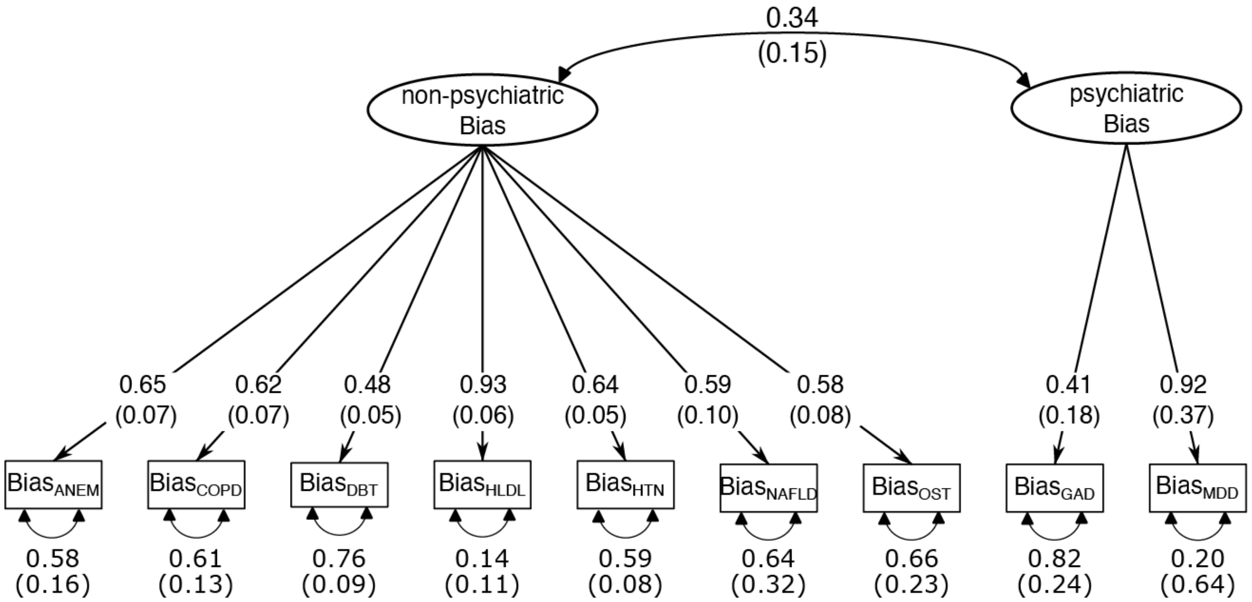

**Supplementary Figure S28.** Psychiatric Bias factor and Non-Psychiatric Bias factor model.

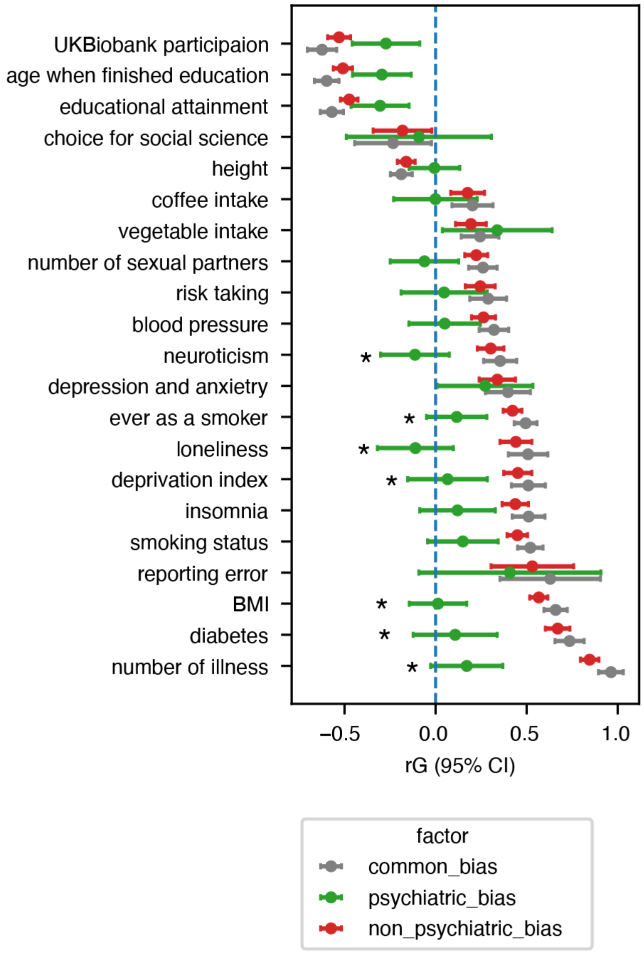

**Supplementary Figure S29.** rG between Psychiatric Bias factor and Non-Psychiatric Bias factor with socioeconomic and behavioral traits. Asterisk (*) indicates a significant difference in rG between the trait and the two types of biases after bonferroni correction. Full results for all traits tested are in **Supplementary Table S10.**

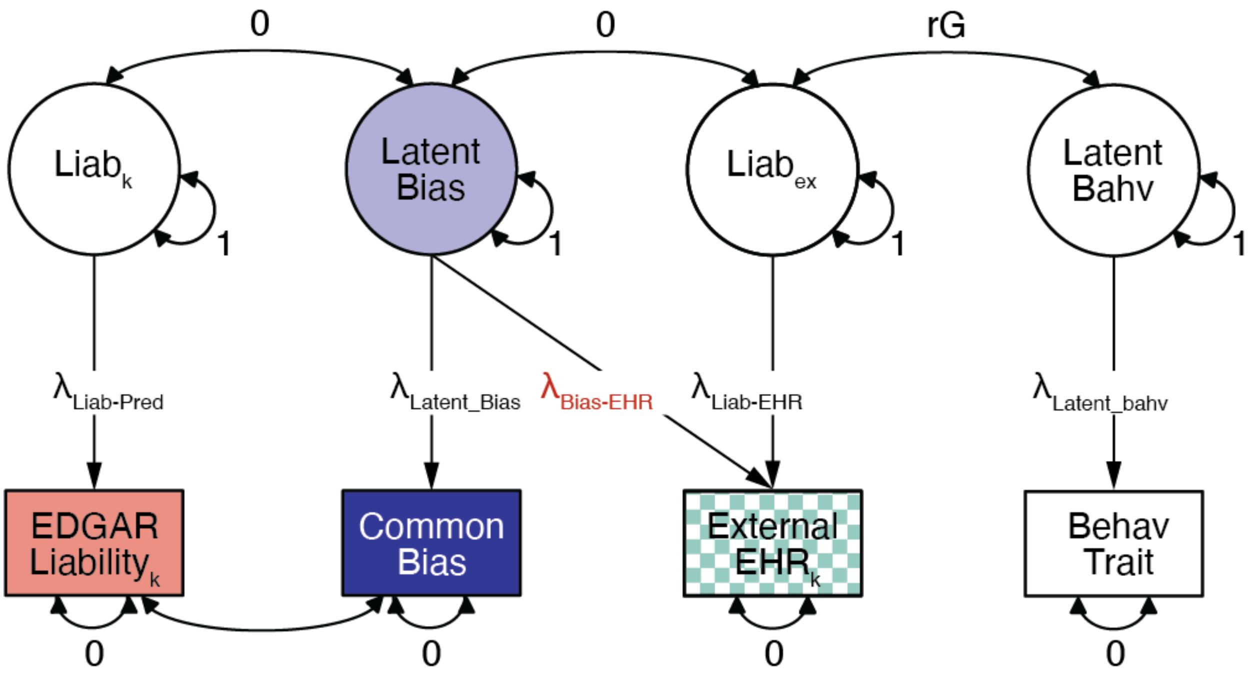

**Supplementary Figure S30.** Illustration of estimating rG with socioeconomic/behavior traits before and after bias removal. We substitute “Behav Tait” with an external deep phenotype when estimating rG with it.
